# How do Covid-19 policy options depend on end-of-year holiday contacts in Mexico City Metropolitan Area? A Modeling Study

**DOI:** 10.1101/2020.12.21.20248597

**Authors:** Fernando Alarid-Escudero, Valeria Gracia, Andrea Luviano, Yadira Peralta, Marissa B. Reitsma, Anneke L. Claypool, Joshua A. Salomon, David M. Studdert, Jason R. Andrews, Jeremy D. Goldhaber-Fiebert, Stanford-CIDE Coronavirus Simulation Model (SC-COSMO) Modeling Consortium

**Author notes:** **Corresponding Author:** Fernando Alarid-Escudero, PhD, Division of Public Administration, Center for Research and Teaching in Economics (CIDE), Circuito Tecnopolo Norte 117, Col. Tecnopolo Pocitos II, Aguascalientes, Aguascalientes, 20313, Mexico. e-mail address; + 5238.

## Abstract

**Background:** With more than 20 million residents, Mexico City Metropolitan Area (MCMA) has the largest number of Covid-19 cases in Mexico and is at risk of exceeding its hospital capacity in late December 2020.

**Methods:** We used SC-COSMO, a dynamic compartmental Covid-19 model, to evaluate scenarios considering combinations of increased contacts during the holiday season, intensification of social distancing, and school reopening. Model parameters were derived from primary data from MCMA, published literature, and calibrated to time-series of incident confirmed cases, deaths, and hospital occupancy. Outcomes included projected confirmed cases and deaths, hospital demand, and magnitude of hospital capacity exceedance.

**Findings:** Following high levels of holiday contacts even with no in-person schooling, we predict that MCMA will have 1·0 million (95% prediction interval 0·5 – 1·7) additional Covid-19 cases between December 7, 2020 and March 7, 2021 and that hospitalizations will peak at 35,000 (14,700 – 67,500) on January 27, 2021, with a >99% chance of exceeding Covid-19-specific capacity (9,667 beds). If holiday contacts can be controlled, MCMA can reopen in-person schools provided social distancing is increased with 0·5 million (0·2 – 1·0) additional cases and hospitalizations peaking at 14,900 (5,600 – 32,000) on January 23, 2021 (77% chance of exceedance).

**Interpretation:** MCMA must substantially increase Covid-19 hospital capacity under all scenarios considered. MCMA’s ability to reopen schools in mid-January 2021 depends on sustaining social distancing and that contacts during the end-of-year holiday were well controlled.

**Funding:** Society for Medical Decision Making, Gordon and Betty Moore Foundation, and Wadhwani Institute for Artificial Intelligence Foundation.

**Research in context:** *Evidence before this study:* As of mid-December 2020, Mexico has the twelfth highest incidence of confirmed cases of Covid-19 worldwide and its epidemic is currently growing. Mexico’s case fatality ratio (CFR) – 9·1% – is the second highest in the world. With more than 20 million residents, Mexico City Metropolitan Area (MCMA) has the highest number and incidence rate of Covid-19 confirmed cases in Mexico and a CFR of 8·1%. MCMA is nearing its current hospital capacity even as it faces the prospect of increased social contacts during the 2020 end-of-year holidays. There is limited Mexico-specific evidence available on epidemic, such as parameters governing time-dependent mortality, hospitalization and transmission. Literature searches required supplementation through primary data analysis and model calibration to support the first realistic model-based Covid-19 policy evaluation for Mexico, which makes this analysis relevant and timely.

*Added value of this study:* Study strengths include the use of detailed primary data provided by MCMA; the Bayesian model calibration to enable evaluation of projections and their uncertainty; and consideration of both epidemic and health system outcomes. The model projects that failure to limit social contacts during the end-of-year holidays will substantially accelerate MCMA’s epidemic (1·0 million (95% prediction interval 0·5 – 1·7) additional cases by early March 2021). Hospitalization demand could reach 35,000 (14,700 – 67,500), with a >99% chance of exceeding current capacity (9,667 beds). Controlling social contacts during the holidays could enable MCMA to reopen in-person schooling without greatly exacerbating the epidemic provided social distancing in both schools and the community were maintained. Under all scenarios and policies, current hospital capacity appears insufficient, highlighting the need for rapid capacity expansion.

*Implications of all the available evidence:* MCMA officials should prioritize rapid hospital capacity expansion. MCMA’s ability to reopen schools in mid-January 2021 depends on sustaining social distancing and that contacts during the end-of-year holiday were well controlled.

## Introduction

The Covid-19 global pandemic reached an estimated 72·9 million confirmed cases and caused 1·6 million deaths by December 17, 2020, with recent incidence rising sharply in low- and middle-income countries (LMICs), especially in Latin America.^1^ Older individuals and those with comorbidities have greater risks of serious health outcomes and death.^2^ Appropriate and timely hospitalization and in-hospital care can mitigate negative health outcomes.^3^ However, even in highly developed countries, rapidly rising case have overwhelmed health systems, reducing their effectiveness.^4^ Hence, governments in LMICs, like Mexico, are deeply concerned about how their less well-resourced healthcare systems will cope with surges in Covid-19 cases.

In mid-December, Mexico had the twelfth highest numbers of confirmed Covid-19 cases worldwide, and the second largest number of Covid-19 deaths in Latin America.^1^ Mexico’s case fatality ratio (CFR) – 9·1% – is the second highest in the world.^5^ With a population of more than 20 million residents, Mexico City Metropolitan Area (MCMA) has the highest number and incidence rate of confirmed Covid-19 cases in Mexico and a CFR of 8·1%.^6^

In the coming months, non-pharmaceutical interventions (NPIs) will remain the primary means of controlling the Covid-19 epidemic in LMICs, even as vaccines are scaled-up globally.^7^ However, because NPIs, especially business and school closures, can be highly disruptive to economic and social wellbeing particularly in settings where many households lack computers and internet connection,^8^ their strictness must be balanced against threats to a functioning healthcare system. Traditional end-of-year holiday festivities, in which many people gather and mix, present particular challenges for Mexico.^9^

To inform Mexico’s decision makers given transmission risks posed by increased end-of-year holiday contacts and the possible health and healthcare impacts of epidemic growth in early 2021, we provide model-based assessments of policy alternatives. The assessments are informed by primary data analyses. In addition to epidemic outcomes, we focus on estimating the risk that MCMA’s hospital system will be saturated by early 2021, and the potential for policies and hospital capacity expansion to mitigate this.

## Methods

### Overview

We implemented a model of MCMA’s Covid-19 epidemic and potential interventions using the Stanford-CIDE Coronavirus Simulation Model (SC-COSMO) framework (appendix pp 13—31). We parameterized the model based on the best-available clinical and epidemiological data from published and publicly available pre-published studies along with primary data on MCMA’s hospitalization and testing infrastructure. We calibrated the model to time-series data on MCMA’s daily confirmed Covid-19 cases, deaths, and hospital occupancy from February 24, 2020 to December 7, 2020. Model calibration determined the joint posterior uncertainty distribution of inputs. The calibrated model projected epidemic and health systems outcomes along with their uncertainty under a range of intervention scenarios from December 7, 2020 to March 7, 2021. Scenarios comprised varying combinations of increased contacts during the end-of-year holiday season followed by intensification of NPIs and re-opening of schools.

### Model structure and assumptions

SC-COSMO is an age-structured, multi-compartment susceptible-exposed-infected-recovered (AS-MC-SEIR) model of SARS-CoV-2 transmission and progression (Figure S8) with realistic demography and contact patterns (household and venue-specific, non-household contacts) enabling finer detail of the interventions considered.^10^ The model is implemented in the R programming language.^11^ SC-COSMO includes both latency and incubation, whose durations are assumed to be gamma distributed (appendix p 20). It incorporates the timing of NPIs (e.g., “social distancing”) and reductions of effective contacts which may differ by age and venue. Forward projections with the SC-COSMO model can compare future scenarios and consider a range of outcomes (e.g., infections, cases, deaths, hospitalizations).

### Scenarios and policies

We used the model to evaluate a range of policies under two scenarios involving heightened levels of social contacts in MCMA during the end-of-year holiday period (December 24, 2020 through January 6, 2021) relative to the level of social contact on December 7, 2020, which is estimated through model calibration. Compared to the calibrated level of reductions in social distancing on December 7, our base case scenario assumes that with less compliance with NPIs, reductions in holiday contacts will be less effective (i.e., we add 0.30 to our calibrated multiplier). In an alternative scenario, we assume that December 7 levels are unchanged under the end-of-year holiday period.

Under each holiday contact scenario, we considered the effect of four different disease control policies followed during the period from December 7, 2020 to March 7, 2021. Policies involved increased compliance with social distancing in the community and in-person school reopening. They included: 1) status quo in which social distancing observed on December 7 again resumes after the holidays with schools remaining closed; 2) increased compliance with community social distancing on January 11, 2021 with schools remaining closed; 3) status quo community social distancing with schools re-opening on January 11, 2021 with status quo in-school contacts; 4) increased compliance with community social distancing with schools re-opening both on January 11, 2021 with reduced in-school contacts.

### Outcomes

Our primary outcomes were time series of incident and cumulative Covid-19 cases, deaths, and hospitalization demand in relation to MCMA hospital capacity. We estimated the effective reproduction number, *R*_*e*_, for March 23, 2020 – the day Mexico implemented national-level NPIs – as well as for all days since then.^12^ We also estimated the likelihood of hospitalization demand exceeding Covid-19-specific capacity over time.

### Data and model inputs

MCMA consists of Mexico City’s counties plus 60 counties of two neighboring Mexican states (Figure S1). The list of counties that form MCMA along with their projected 2020 population is shown in Table S1. Overall demographic data on MCMA including its age-structure and age-specific background mortality rates were derived from official statistics (Table 1).^14^ We collapsed ages into eight groups that reflected likely patterns of exposures (e.g., school age children, retirees, etc.).

**Table 1.** Demographic, health system capacity and Covid-19 outcome data for Mexico City Metropolitan Area (MCMA)

|  | Value | Source |
| --- | --- | --- |
| **Demography** |  |  |
| Total population in the state of MCMA | **21,942,666** | CONAPO <sup>14</sup> |
| Population density (population/mi<sup>2</sup>) | **53,339** | CONAPO<sup>14</sup> and INEGI<sup>30</sup> |
| **Health system Covid-19 capacity as of December 7, 2020** |  |  |
| Total hospital beds | **9,667** | Digital Agency for Public Innovation<sup>13</sup> |
| Beds with ventilators | **2,659** | Digital Agency for Public Innovation<sup>13</sup> |
| **Covid-19 outcomes as of December 7, 2020** |  |  |
| Cases | **344,028** | Ministry of Health of Mexico<sup>6</sup> |
| Cumulative case rate (per 100,000) | **1,167** | Author's calculation |
| Deaths | **27,733** | Ministry of Health of Mexico<sup>6</sup> |
| Cumulative death rate (per 100,000) | **126** | Author's calculation |
| Hospitalized patients | **68,225** | Ministry of Health of Mexico<sup>6</sup> |
| Hospitalized patients requiring ventilator | **12,458** | Ministry of Health of Mexico<sup>6</sup> |

We compiled publicly available data from Mexico’s Ministry of Health on all detected cases and deaths in MCMA from February 24 through December 7, 2020.^6^ We used these data to compute daily incident and cumulative confirmed cases and deaths, and to estimate time-varying case fatality rates with proportional hazard models that included splines on calendar time.

We received daily updates from MCMA’s Digital Agency for Public Innovation^13^ on hospital inpatient census of severe-acute respiratory infection (SARI) beds with and without ventilator occupancy, as well as current hospital capacity, which has expanded over time. We estimated time-varying hospital length of stay for Covid-19 patients, stratified by whether or not they required a ventilator via model calibration.

Literature reviews provided Covid-19-specific epidemiologic parameters. Latent and incubation periods were assumed to follow a gamma distribution (appendix p 20).^15–17^ Notably, the probability of hospitalization and death among cases are not derived from the literature; they are estimated from the primary data, as described above.

### Model calibration and uncertainty analysis

We used Bayesian methods to calibrate 11 model parameters that could not be directly estimated from data. The parameters concerned transmissibility in the community and in the household, time-varying effects of MCMA’s NPIs expressed as proportionate reduction of pre-epidemic levels of daily effective contacts, and time-varying rates of case detection. Calibration inferred values for model parameters by matching modeled outcomes to daily incident confirmed Covid-19 cases (i.e., calibration targets) from February 24 to December 7, 2020. Comparison of modeled outcomes and empirical data used a likelihood function, which we constructed by assuming that targets follow negative binomial distributions with means given by the model-predicted outputs and a dispersion equal to one, to account for potential overdispersion in the target data.^18^ We defined uniform prior distributions for all calibrated parameters with ranges based on existing evidence, epidemic theory, and plausibility (Table 2). Calibration resulted in an estimate of the joint posterior uncertainty distribution for the model parameters.

**Table 2.**
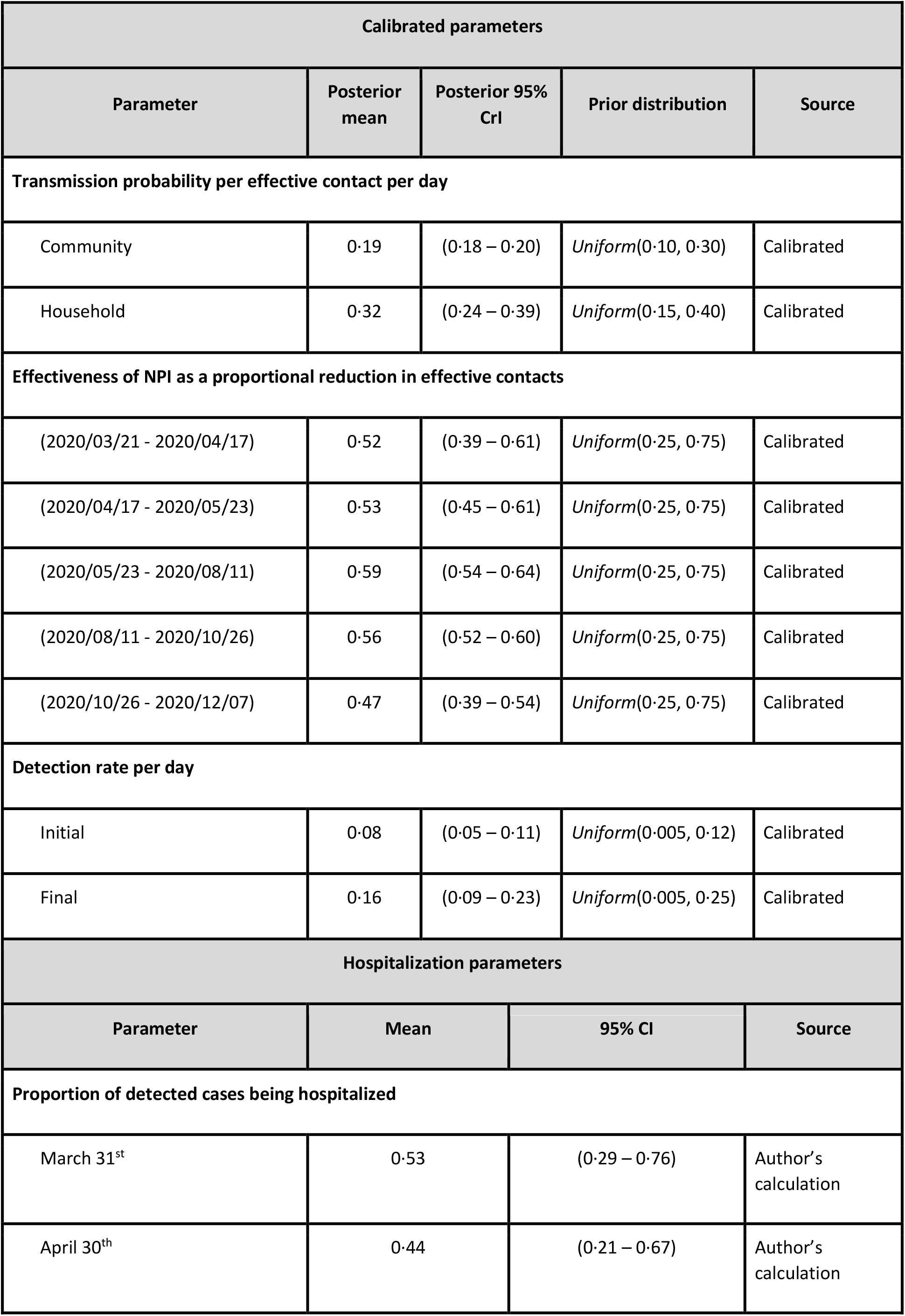
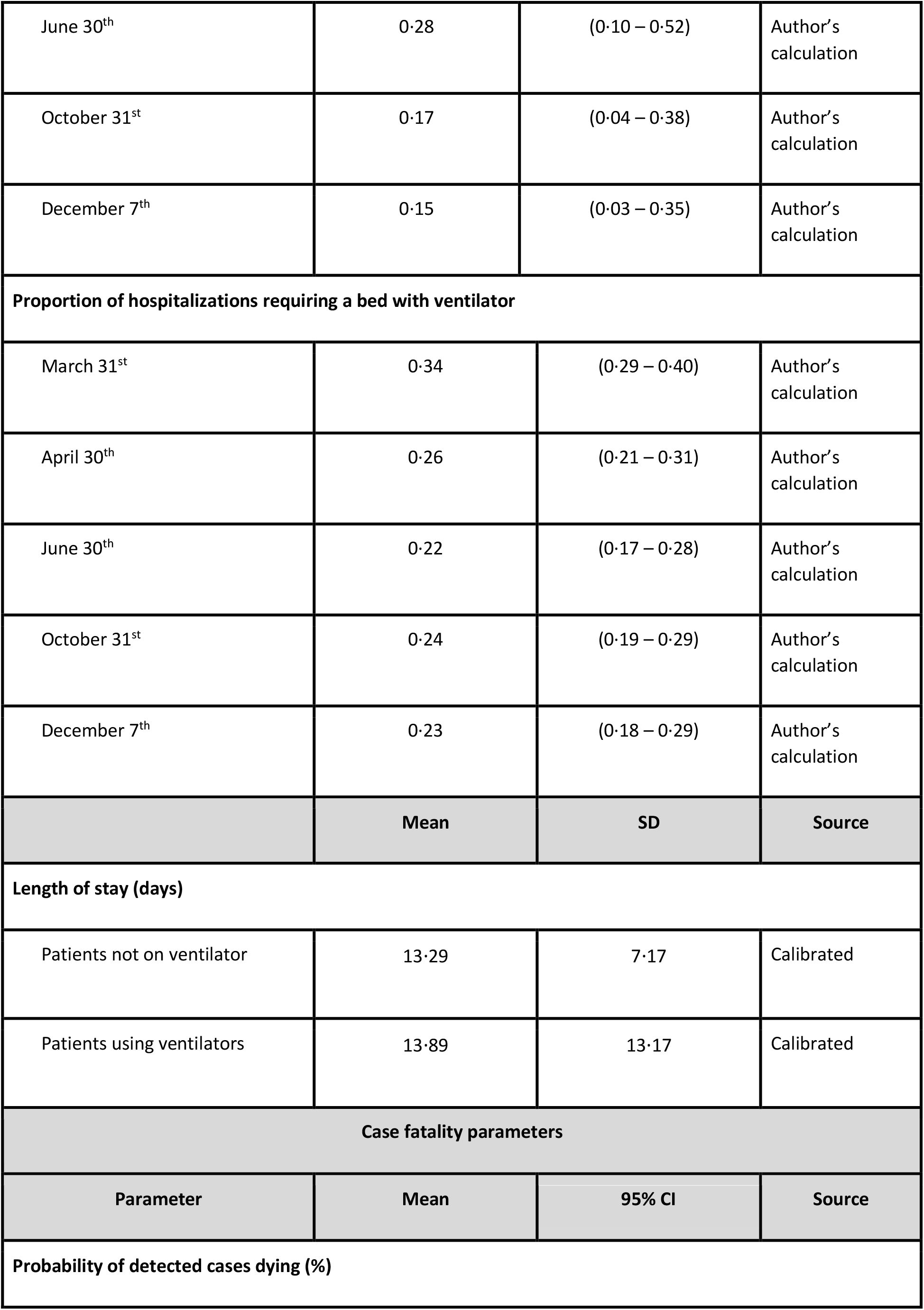
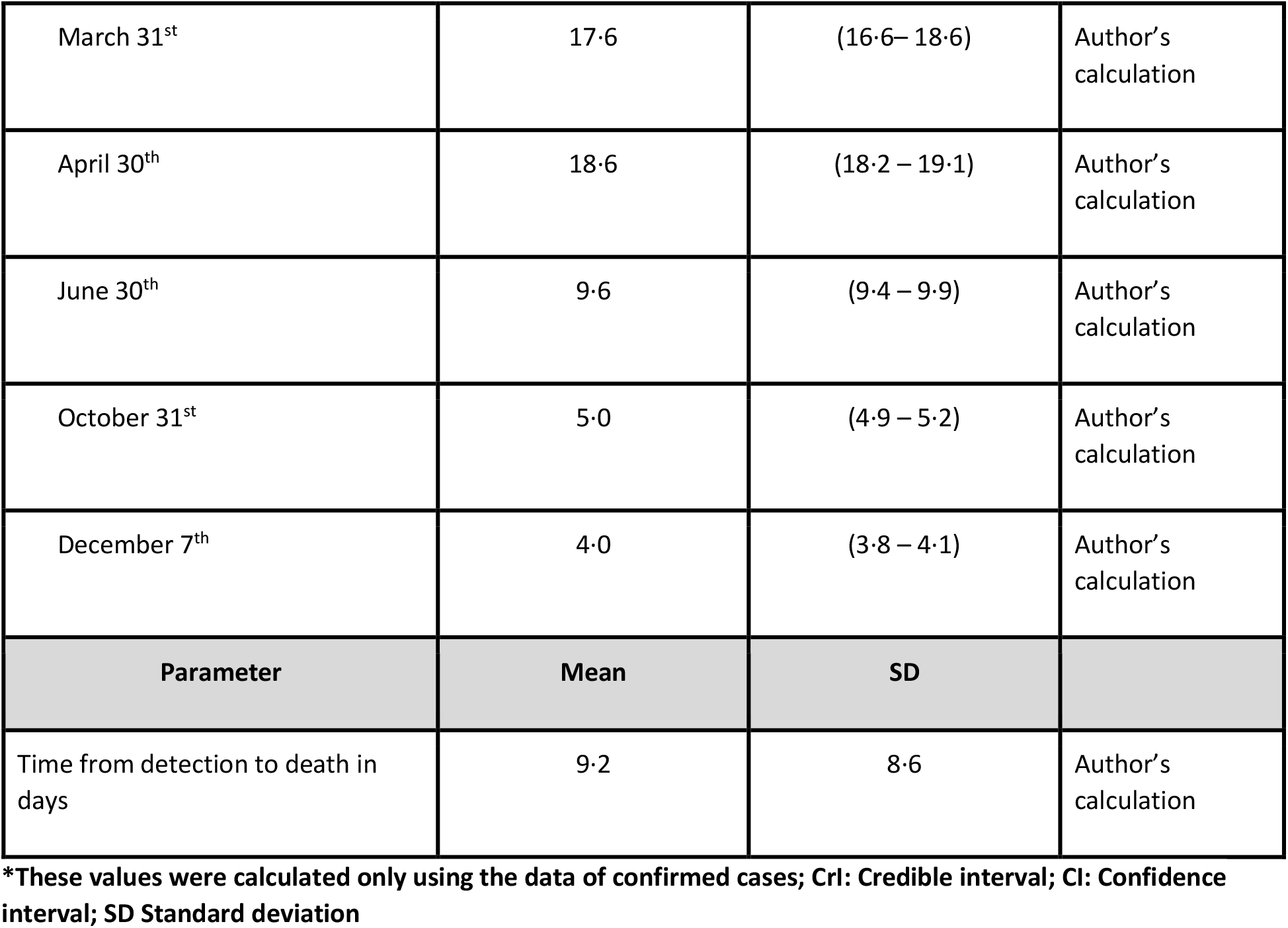
Model input parameters

| Calibrated parameters |  |  |  |  |
| --- | --- | --- | --- | --- |
| Parameter | Posterior mean | Posterior 95% CrI | Prior distribution | Source |
| Transmission probability per effective contact per day |  |  |  |  |
| Community | 0.19 | (0.18 – 0.20) | Uniform(0.10, 0.30) | Calibrated |
| Household | 0.32 | (0.24 – 0.39) | Uniform(0.15, 0.40) | Calibrated |
| Effectiveness of NPI as a proportional reduction in effective contacts |  |  |  |  |
| (2020/03/21 - 2020/04/17) | 0.52 | (0.39 – 0.61) | Uniform(0.25, 0.75) | Calibrated |
| (2020/04/17 - 2020/05/23) | 0.53 | (0.45 – 0.61) | Uniform(0.25, 0.75) | Calibrated |
| (2020/05/23 - 2020/08/11) | 0.59 | (0.54 – 0.64) | Uniform(0.25, 0.75) | Calibrated |
| (2020/08/11 - 2020/10/26) | 0.56 | (0.52 – 0.60) | Uniform(0.25, 0.75) | Calibrated |
| (2020/10/26 - 2020/12/07) | 0.47 | (0.39 – 0.54) | Uniform(0.25, 0.75) | Calibrated |
| Detection rate per day |  |  |  |  |
| Initial | 0.08 | (0.05 – 0.11) | Uniform(0.005, 0.12) | Calibrated |
| Final | 0.16 | (0.09 – 0.23) | Uniform(0.005, 0.25) | Calibrated |
| Hospitalization parameters |  |  |  |  |
| Parameter | Mean | 95% CI | Source |  |
| Proportion of detected cases being hospitalized |  |  |  |  |
| March 31<sup>st</sup> | 0.53 | (0.29 – 0.76) | Author's calculation |  |
| April 30<sup>th</sup> | 0.44 | (0.21 – 0.67) | Author's calculation |  |

|  |  |  |  |
| --- | --- | --- | --- |
| June 30<sup>th</sup> | 0·28 | (0·10 – 0·52) | Author's calculation |
| October 31<sup>st</sup> | 0·17 | (0·04 – 0·38) | Author's calculation |
| December 7<sup>th</sup> | 0·15 | (0·03 – 0·35) | Author's calculation |
| **Proportion of hospitalizations requiring a bed with ventilator** |  |  |  |
| March 31<sup>st</sup> | 0·34 | (0·29 – 0·40) | Author's calculation |
| April 30<sup>th</sup> | 0·26 | (0·21 – 0·31) | Author's calculation |
| June 30<sup>th</sup> | 0·22 | (0·17 – 0·28) | Author's calculation |
| October 31<sup>st</sup> | 0·24 | (0·19 – 0·29) | Author's calculation |
| December 7<sup>th</sup> | 0·23 | (0·18 – 0·29) | Author's calculation |
|  | **Mean** | **SD** | **Source** |
| **Length of stay (days)** |  |  |  |
| Patients not on ventilator | 13·29 | 7·17 | Calibrated |
| Patients using ventilators | 13·89 | 13·17 | Calibrated |
| **Case fatality parameters** |  |  |  |
| **Parameter** | **Mean** | **95% CI** | **Source** |
| **Probability of detected cases dying (%)** |  |  |  |
| March 31<sup>st</sup> | 17·6 | (16·6– 18·6) | Author's calculation |
| April 30<sup>th</sup> | 18·6 | (18·2 – 19·1) | Author's calculation |
| June 30<sup>th</sup> | 9·6 | (9·4 – 9·9) | Author's calculation |
| October 31<sup>st</sup> | 5·0 | (4·9 – 5·2) | Author's calculation |
| December 7<sup>th</sup> | 4·0 | (3·8 – 4·1) | Author's calculation |
| **Parameter** | **Mean** | **SD** |  |
| Time from detection to death in days | 9·2 | 8·6 | Author's calculation |

\*These values were calculated only using the data of confirmed cases; CrI: Credible interval; CI: Confidence interval; SD Standard deviation

To conduct the Bayesian calibration, we used the incremental mixture importance sampling (IMIS) algorithm,^19^ which has been previously used to calibrate health policy models.^20^ Briefly, we sampled 10,000 parameter sets from our priors in the first stage followed by 1,000 samples in each of the consecutive 45 updated sampling stages. This procedure yielded a posterior distribution from which we obtained 1,000 samples used for our projections and analyses. The marginal posterior distributions and pairwise comparisons are shown in Figures S2 and S3.

For all outcome measures, we accounted for model input parameter uncertainty by randomly sampling from the joint posterior distribution obtained from the Bayesian calibration. We used 1,000 parameter sets sampled from the posterior distribution to generate all primary outcomes for all scenarios and policies with 95% posterior model-prediction intervals (PI) for each outcome from the 2·5^th^ and 97·5^th^ percentiles of the projected values.

## Results

### MCMA’s epidemic to date

The Covid-19 epidemic in MCMA has involved substantial burdens of cases, hospitalizations, and deaths, which the SC-COSMO model replicates (Figures 1 and 2). Case rates rose from mid-March through late May, remained high through mid-October, and have steadily increased since then (Figure 1). Trends in deaths and hospitalizations have followed the same general pattern. By December 7, 2020, MCMA had experienced 344,028 confirmed Covid-19 cases and 27,733 deaths (Table 1), which represent cumulative incident and mortality rates of 1,167 and 126 per 100,000 population, respectively. Among confirmed cases, 68,225 (20%) involved hospitalizations and 12,458 (18%) involved ventilator hospitalizations. Patients died in 29% of non-ventilator hospitalizations and 80% of ventilator hospitalizations. The average length of stay in non-ventilator hospitalizations was 13·29 days (standard deviation [SD] of 7·17); the average length of stay in ventilator hospitalizations was 13·89 days (SD of 13·17).

**Figure 1.**
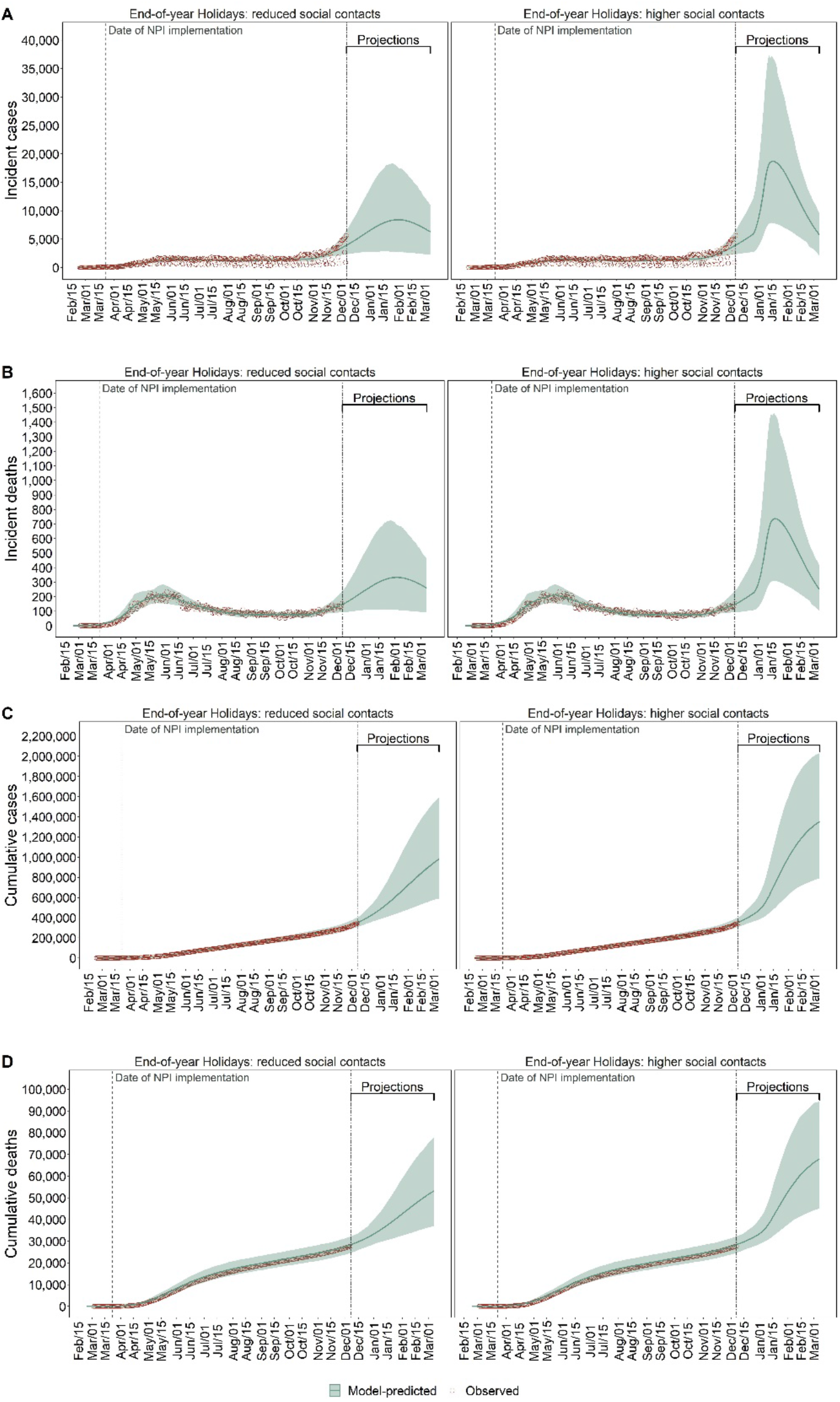
Observed (red dots) and model-predicted (green lines) Covid-19 incident detected cases (A), deaths (B), cumulative cases (C), and deaths (D) in MCMA between February 24, 2020 and March 7, 2021. Left column plots assume compliance with social distancing during end-of-year holiday period. Right column plots assume substantially less compliance with social distancing during the end-of-year holiday period. Double-dashed vertical line represents the last day used for calibration. The green shaded area shows the 95% posterior model-predictive interval of the outcomes and the green lines show the posterior model-predicted mean based on 1,000 simulations using samples from posterior distribution.

**Figure 2.**
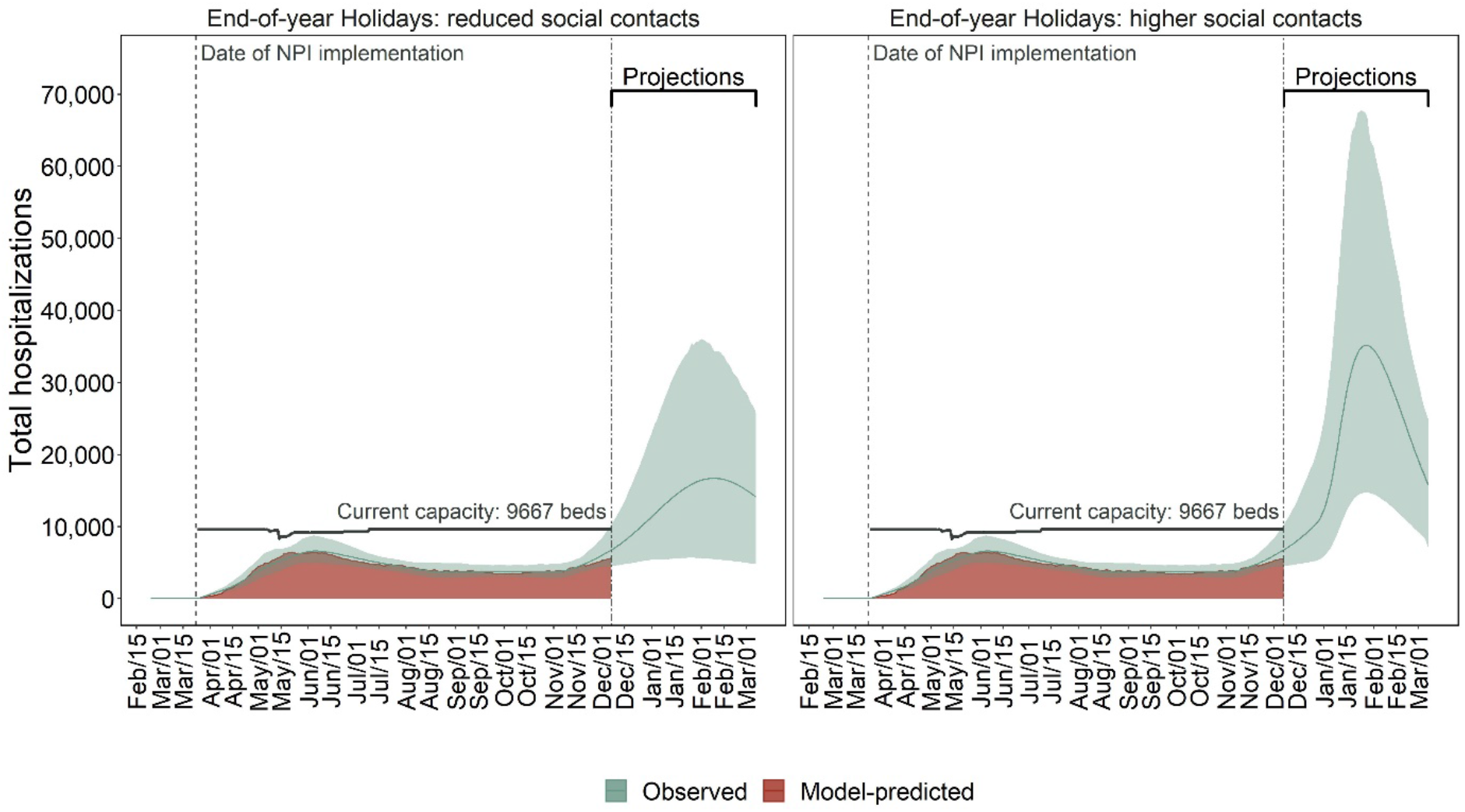
Observed (red area) and model-predicted (green lines) total hospital occupancy and demand in MCMA between February 24, 2020 and March 7, 2021. Left plot assumes compliance with social distancing during end-of-year holiday period. Right plot assumes substantially less compliance with social distancing during the end-of-year holiday period. Double-dashed vertical line represents the last day used for calibration. The green shaded area shows the 95% posterior model-predictive interval of the outcomes and the colored lines show the posterior model-predicted mean based on 1,000 simulations using samples from posterior distribution. The horizontal black lines show total Covid-19-specific hospital capacity.

On March 17, 2020, *R*_*e*_ for Covid-19 in MCMA was 2·23 (95% PI: 2·12 – 2·35) and decreased to 1·37 (1·28 – 1·47) immediately after implementation of NPIs and was 1·10 (1·03 – 1·18) by December 7, 2020 (Figure S4). This implies that given the estimated *R*_*e*_ in the early phases of the epidemic and if pre-pandemic contact patterns had not changed early on, MCMA would have experienced a bigger Covid-19 epidemic than that observed.

Effective contact rates were substantially decreased after MCMA’s NPIs were originally implemented but have subsequently increased. Specifically, the calibrated model estimated that effective contacts were 52% (95% PI: 39 – 61) lower than pre-pandemic levels in late March 2020 (Table 2 and Figures S2 and S3) but only 47% lower (40 - 54) in early December. Cumulatively, 7% (5 – 10) of the MCMA population – representing 1·5 million people (1·5 – 2.2) – had previously been infected by December 7, 2020 (see Figure S5), 24% (15 - 32) of whom were detected as cases (see Figure S6).

### Contact patterns and epidemic risks during the end-of-year holidays

The trajectory of MCMA’s epidemic from late December through mid-January 2021 depends heavily on the extent to which gatherings that traditionally take place in Mexico during the end-of-year holidays occur and cause effective contact rates to rise. Our base case assumption of increased contacts during the holidays projects a peak of 18,708 (95% PI 7,821 – 36,874) daily incident cases and 737 (308 – 1,450) deaths on mid-January 2021 (Figure 1). However, if compliance with social distancing reduces contacts compared to previous years in this holiday period, daily incident cases and deaths could have a lower peak at 8,420 (2,749 – 17,668) and 333 (109 – 696), respectively, on early-February 2021 (Figures 1).

Demand for hospitalization is likely to exceed Covid-19-specific hospital capacity by early 2021, even if end-of-year holiday contacts are reduced, although these contacts will strongly determine the extent to which capacity is exceeded and the duration of exceedance. Peak demand on January 27, 2021 is projected to be 35,000 (14,700 – 67,500) with high levels of holiday contacts and 16,711 (5,494 – 34,372) on February 9, 2021 if holiday contacts are reduced (Figure 2); both far exceed MCMA’s current capacity of 9,667 hospital beds. The likelihood of exceeding hospital capacity by these dates is >99% with high levels of holiday contacts versus 80% with low levels of contacts (Figure 5).

**Figure 5.**
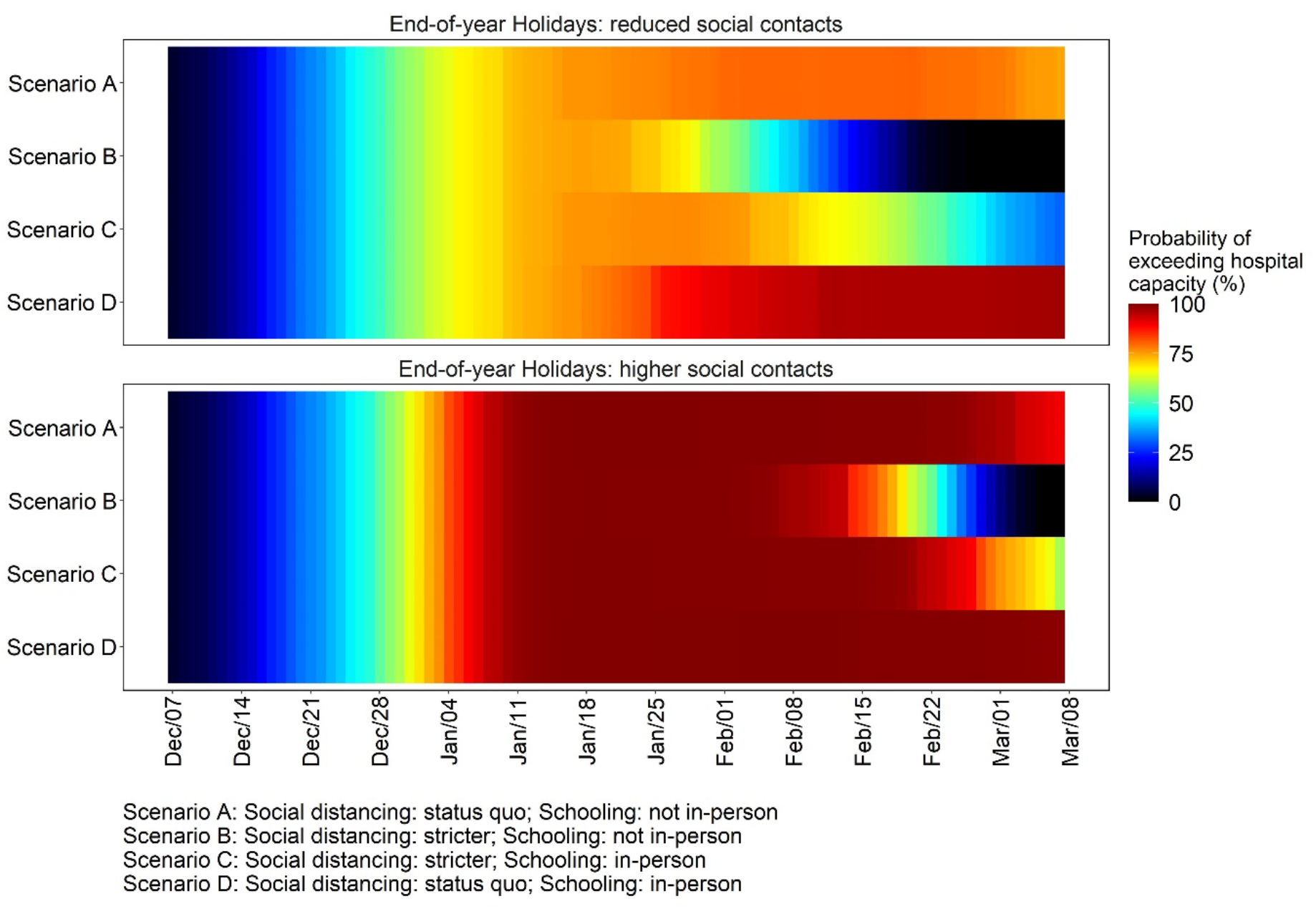
Daily estimated likelihood of hospitalization demand exceeding Covid-19-specific capacity in MCMA between December 7, 20201 and March 7, 2021 by levels of compliance with social distancing during end-of-year holiday period. Top panel assumes compliance with social distancing during end-of-year holiday period. Bottom panel assumes substantially less compliance with social distancing during the end-of-year holiday period.

### Policy analysis without social distancing compliance during the 2020 end-of-year holiday period

NPI policies and compliance from mid-January through early March 2021 will play important roles in mitigating adverse health outcomes and the speed and extent to which hospitalization demands exceeds capacity. The following policy comparisons assume that with less compliance with NPIs, holiday contacts are only 17% (95% PI 9 – 24) lower than pre-pandemic levels.

#### Social distancing: status quo; Schooling: not in-person

If mid-December levels of contacts resume after the holidays and in-person schools remained closed (i.e., the status-quo), we estimate the following for March 7, 2021: 5,787 (95% PI 2,142 – 9,582) incident daily cases and 253 (105 – 418) incident daily deaths (Figure 3); *R*_*e*_ of 0·84 (0·73 – 0·93), indicating a declining but still substantial epidemic (Figure S4); hospital demand of 15,794 (7,124 – 24,908) (Figure 4), and a 90% likelihood of exceeding Covid-19-specific capacity (Figure 5).

**Figure 3.**
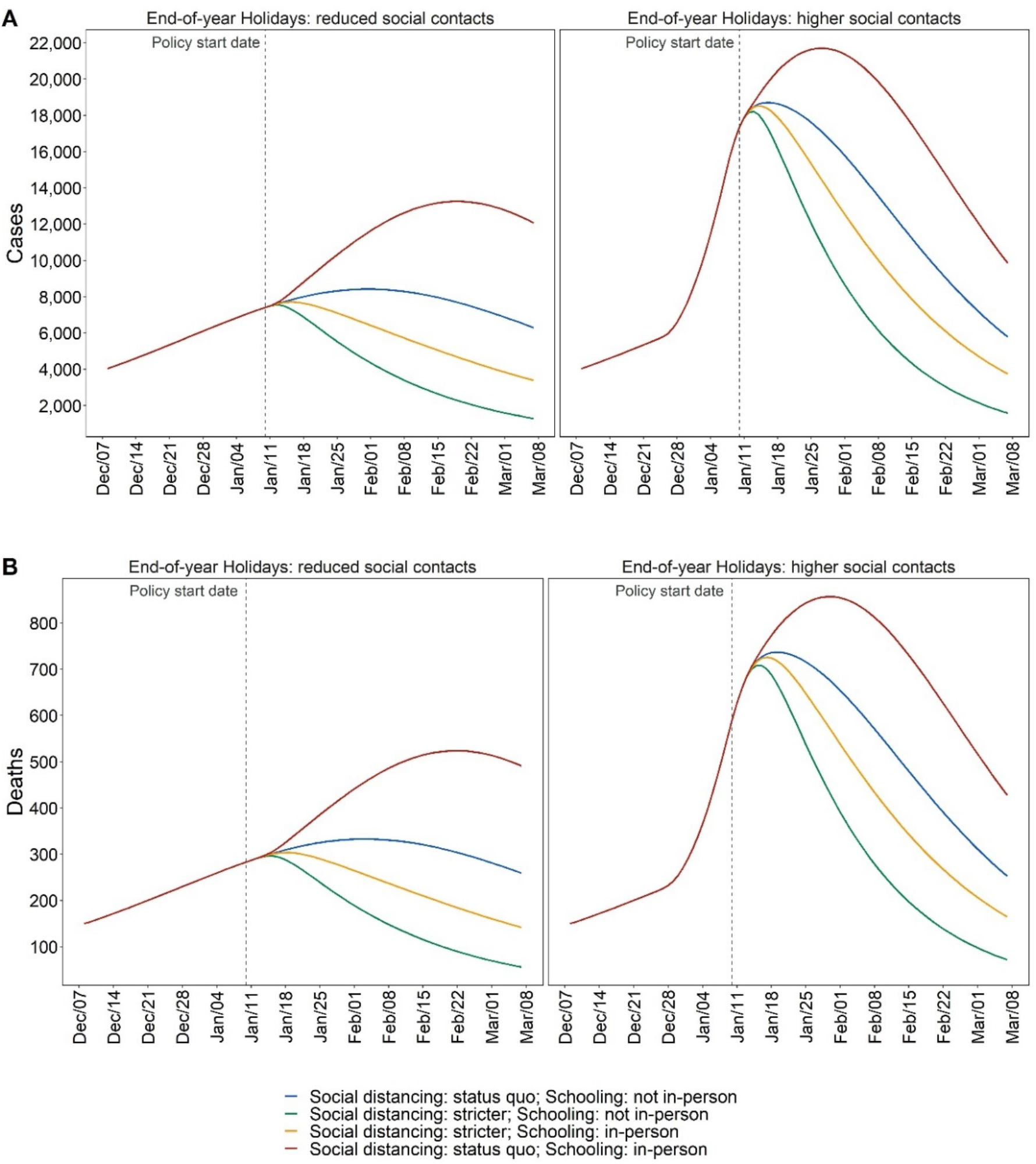
Estimated model-predicted daily incident cases (A) and deaths (B) by scenario in MCMA between December 7, 2020 to March 7, 2021. Left column plots assume compliance with social distancing during end-of-year holiday period. Right column plots assume substantially less compliance with social distancing during the end-of-year holiday period. Vertical dashed line represents the day of policy implementations.

**Figure 4.**
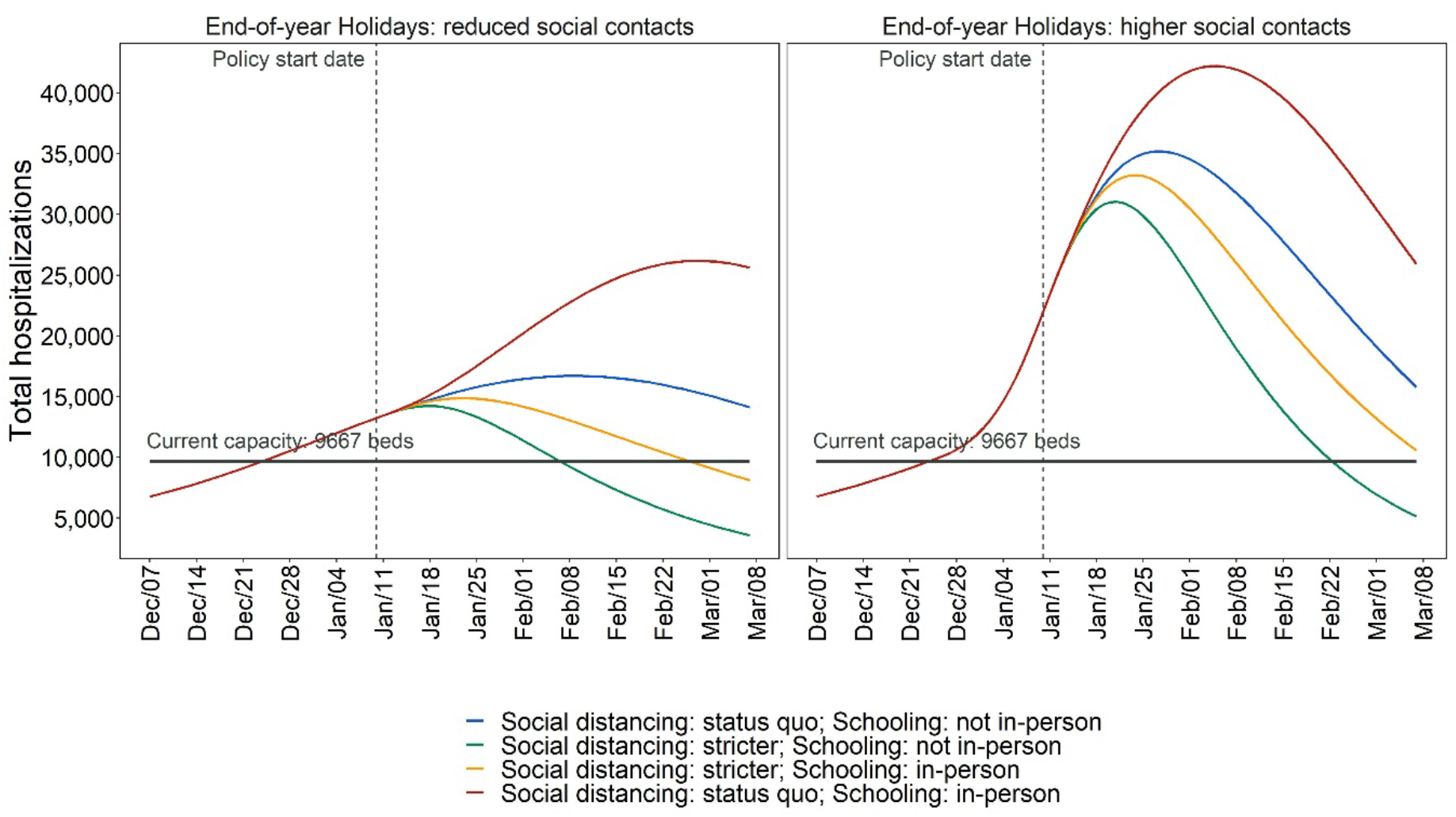
Estimated model-predicted daily hospitalization demand in MCMA between December 7, 2020 and March 7, 2021. Left column plots assume compliance with social distancing during end-of-year holiday period. Right column plots assume substantially less compliance with social distancing during the end-of-year holiday period.

#### Social distancing: stricter; Schooling: not in-person

However, if on January 10, 2021, social distancing was intensified relative to December 7 levels resulting in contacts being 59% (95% PI 54 – 64) lower than pre-pandemic levels, and in-person schools remained closed, these outcomes would be substantially better. By March 7, we estimate 1,584 (441 – 2,880) incident daily cases and 72 (22 – 128) incident daily deaths (Figure 3); *R*_*e*_ of 0·78 (0·67 – 0·86) (Figure S4); hospital demand at 5,147 (2,007 – 8,287) (Figure 4), with a <1% likelihood of exceeding capacity (Figure 5).

### In-person school reopening in early 2021

Reopening in-person schooling is a high priority given the negative societal impacts of these closures. However, epidemic outcomes depend on how reopening is implemented and how much social distancing can be achieved both in schools and other community venues.

#### Social distancing: status quo; Schooling: in-person

Resumption of in-person schooling without reductions in contacts would result in appreciably greater epidemic growth. By March 7, incident daily cases and deaths would be 9,851 (3,406– 16,330) and 428 (156 – 692) respectively (Figure 3); *R*_*e*_ would be 0·84 (95% PI 0·72 – 0·95) (Figure S4); and hospital demand would be 25,908 (11,984 – 40,405) with the likelihood of exceeding capacity at >99% (Figures 4 and 5).

#### Social distancing: stricter; Schooling: in-person

However, if in-person school resumes with contacts reduced substantially below mid-December levels in both schools and the community, then epidemic and hospitalization outcomes would be slightly better than the status quo without school reopening. By March 7, incident daily cases and deaths would be 3,742 (95% PI 1,078 – 6,698) and 150 (92– 246) respectively (Figure 3); *R*_*e*_ would be 0·83 (95% PI 0·71 – 0·92) (Figure S4); hospitalization demand would be 10,593 (4,134 – 17,013) with the likelihood of exceeding capacity at 59% (Figures 4 and 5).

### Policy analysis with social distancing compliance during the 2020 end-of-year holiday period

Results of the policy comparisons under a scenario that assumes there are not high levels of end-of-year holiday contacts are consistent with those presented above for the base case. While our main health and hospital outcomes are generally less extreme owing to lower transmission in the period between December 24, 2020 and January 6, 2021, the rank ordering of policies by efficacy does not change (Figures 1-5 and S4). Importantly, if social distancing compliance were achieved during the end-of-year holiday period, in-person school reopening with appropriate social distancing would be feasible in mid-January 2021 without sparking substantial additional epidemic growth, although hospitalization capacity would be exceeded. Importantly, the only scenario in which Covid-19-specific hospital capacity is not exceeded is under stricter community social distancing.

## Discussion

For Mexico City Metropolitan Area’s population of 20 million people, we estimated the epidemic and hospital system effects of resuming in-person schooling in early 2021 and how these effects depend upon the level of end-of-year holiday contacts. Regardless of the level of social distancing MCMA residents are able to achieve during the holidays, hospital demand is very likely to exceed current capacity unless resources are quickly expanded. We found that high levels of end-of-year holiday contacts greatly exacerbate cases and deaths, with lasting effects through early March 2021, and that these effects could be substantially attenuated by greater social distancing during the end-of-year holiday period. Without improved social distancing during the holidays, reopening in-person schools, even with augmented social distancing, results in appreciable epidemic growth. Thus, we conclude that the feasibility of re-opening in-person schooling in the new year depends on reducing mixing and social contacts during the holidays.

While we find that MCMA is expected to exceed hospital capacity as cases continue to rise across scenarios and policies, the timing and magnitude of exceedance differs by scenario. Nonetheless, to meet the surge in hospital demand expected even under optimistic scenarios, MCMA may have to increase Covid-19-specific capacity by at least 4,500 beds.

Reopening in-person schooling is a high priority, and our findings suggest that provided social distancing can be maintained both at schools and in the community, reopening may be possible without substantial additional impact to epidemic and health system outcomes. However, if social distancing cannot be complied with or enforced, school reopening could increase confirmed cases by 410,000 compared to reopening with strict social distancing. Furthermore, we found that the extent of transmission during the end-of-year policies has an important effect on the feasibility of reopening schools without sparking additional epidemic growth, which is consistent with other findings.^21^

Our results are in line with those from previous modeling studies of NPIs in general and compliance with social distancing in particular.^3,22,23^ Briefly, strengthening both has tremendous potential to reduce epidemic transmission, as does closure of in-person school. But these policies can be highly disruptive, trigger other economic and social costs, and are increasingly provoking backlashes among frustrated and weary communities throughout the world. Our findings underscore a theme evident in similar studies: policy decisions about reopening various venues and institutions are interrelated in their effects and must be considered as part of the trade-offs that include health, economic, and social outcomes.

Our analysis has several limitations. First, while our model is stratified by age to account for differential mixing, it does not include differential transmission by younger people.^24^ Some studies find that younger children may transmit less than teens and adults.^22,24^ If children are differentially less likely to transmit then, at least for primary schools, our results may understate the possibility of resuming in-person schooling with social distancing without exacerbating the epidemic and could be viewed as conservative. Second, our analysis does not account for vaccination. However, the time periods we focus on precede plausible mass vaccination, given current expectations regarding vaccine roll-out in Mexico.^25^ Third, we purposefully focus on the health and health systems impacts of policy alternatives needed in the short-term and do not conduct a cost-effectiveness analysis (CEA) over a lifetime horizon. Hence, we do not quantify the full costs and health outcomes associated with missing school^26^ or work.^27^ Findings from our analysis will be useful inputs for a wider economic evaluation of policy alternatives in future research.

Our study has several strengths. First, we use the SC-COSMO model, which is a dynamic transmission model that accounts for realistic contact patterns based on adjusted population density^28^ and both community and household transmission.^10^ The SC-COSMO framework enables quantification and propagation of uncertainty to generate probabilistic projections – not only producing estimates of expected outcomes, but also allowing assessment of the likelihood and magnitude of extreme events like exceeding hospital capacity under different scenarios.^29^ We use comprehensive data on cases, deaths and hospitalizations to estimate the parameters of the model, which allows us to accurately represent the epidemic dynamics in MCMA. Additionally, we have information on current hospital capacity in the city that allows us to determine when and how likely it is to exceed Covid-19-specific hospital capacity under different scenarios.

As MCMA’s Covid-19 epidemic continues to evolve, there is a high probability that the area’s hospital capacity will be outstripped by early January 2021, especially if contacts during the end-of-year holidays cannot be substantially reduced. As resumption of in-person school is a major priority, it is important to ensure that NPI measures are instituted in schools and that the unavoidable increases in contacts that school reopenings will trigger are offset by more effective social distancing in the community. Even if schools are not reopened and social distancing in the community improves, there is an urgent need for MCMA to increase its hospital capacity. Finally, our findings highlight the importance of simulation modeling-based policy analysis as a tool to support timely decision making.

## Data Availability

Analysis code and data are available from the corresponding author upon request.

## Contributors

FAE contributed to the conceptualization, data curation, formal analysis, funding acquisition, investigation, methodology, project administration, resources, software, supervision, validation, visualization, and writing of the original draft and of reviewing and editing; JGF contributed to the conceptualization, formal analysis, funding acquisition, investigation, methodology, resources, validation, and writing of the original draft and of reviewing and editing; VG contributed to the data curation, formal analysis, validation, visualization, and writing of the original draft and of reviewing and editing; AL contributed to the data curation, formal analysis, validation, visualization, and writing of the original draft and of reviewing and editing; YP contributed to the data curation, formal analysis, funding acquisition, investigation, methodology, project administration, resources, software, supervision, validation, and reviewing and editing of the original draft; MBR contributed to the methodology, software, and writing of the original draft and of reviewing and editing; ALC contributed to the methodology, software, and writing of the original draft and of reviewing and editing; DMS contributed to the investigation, and writing of the original draft and of reviewing and editing; JAS contributed to the conceptualization, investigation, and writing of the original draft and of reviewing and editing; JRA contributed to the investigation, methodology, and writing of the original draft and of reviewing and editing.

## Data sharing statement

Analysis code and data are available from the corresponding author upon request.

## Acknowledgements

Drs. Alarid-Escudero and Peralta were supported by a grant from the Society for Medical Decision Making (SMDM) funded by the Gordon and Betty Moore Foundation and a grant from Open Society Foundations (OSF). Dr. Salomon was supported by funding from the Centers for Disease Control and Prevention though the Council of State and Territorial Epidemiologists (NU38OT000297-02) and the National Institute on Drug Abuse (3R37DA01561217S1). Our work on epidemic control of Covid-19 relevant for low- and middle-income countries was supported in part by a gift from the Wadhwani Institute for Artificial Intelligence Foundation. The funders had no role in study design, data collection, data analysis, data interpretation, writing of the article, or the decision to submit for publication. All authors had full access to all the data in the study and were responsible for the decision to submit the article for publication. We have not been paid to write this article by any pharmaceutical company or other agency. The authors are grateful for feedback received for development and model use during the Covid-19 epidemic from researchers, policymakers, and stakeholders.

