## appendix for "How do Covid-19 policy options depend on end-of-year holiday contacts in Mexico City Metropolitan Area? A Modeling Study"

#### For

**Contents**

1. Mexico City Metropolitan Area (MCMA)
2. Calibrated posterior distributions
3. Supplementary results
4. Stanford-CIDE Coronavirus Simulation Model (SC-COSMO) framework

#### 1. Mexico City Metropolitan Area (MCMA)

**Table S1.** Mexico City Metropolitan Area: projected population in 2020.

| Municipality | Total |
|---|---|
| **Hidalgo State** | |
| Tizayuca | 137,165 |
| **Mexico City** | |
| Alvaro Obregon | 755,537 |
| Azcapotzalco | 408,441 |
| Benito Juarez | 433,708 |
| Coyoacan | 621,952 |
| Cuajimalpa de Morelos | 199,809 |
| Cuauhtemoc | 548,606 |
| Gustavo A. Madero | 1,176,967 |
| Iztacalco | 393,821 |
| Iztapalapa | 1,815,551 |
| La Magdalena Contreras | 245,147 |
| Miguel Hidalgo | 379,624 |
| Milpa Alta | 139,371 |
| Tlahuac | 366,586 |
| Tlalpan | 682,234 |
| Venustiano Carranza | 433,231 |
| Xochimilco | 418,060 |
| **State of Mexico** | |
| Acolman | 186,256 |
| Amecameca | 54,548 |
| Apaxco | 31,576 |
| Atenco | 70,016 |
| Atizapan de Zaragoza | 557,108 |
| Atlautla | 32,674 |
| Axapusco | 30,040 |
| Ayapango | 11,081 |
| Chalco | 397,344 |
| Chiautla | 31,803 |
| Chicoloapan | 226,911 |
| Chiconcuac | 27,570 |

| Municipality | Total |
|---|---|
| Chimalhuacan | 720,207 |
| Coacalco de Berriozabal | 310,743 |
| Cocotitlan | 15,387 |
| Coyotepec | 44,201 |
| Cuautitlan | 175,004 |
| Cuautitlan Izcalli | 577,190 |
| Ecatepec de Morelos | 1,707,754 |
| Ecatzingo | 10,090 |
| Huehuetoca | 147,326 |
| Hueypoxtla | 46,742 |
| Huixquilucan | 290,231 |
| Isidro Fabela | 12,512 |
| Ixtapaluca | 551,034 |
| Jaltenco | 29,179 |
| Jilotzingo | 20,713 |
| Juchitepec | 27,241 |
| La Paz | 309,596 |
| Melchor Ocampo | 61,172 |
| Naucalpan de Juarez | 910,187 |
| Nextlalpan | 43,640 |
| Nezahualcoyotl | 1,135,786 |
| Nicolas Romero | 441,064 |
| Nopaltepec | 9,753 |
| Otumba | 38,186 |
| Ozumba | 31,154 |
| Papalotla | 4,367 |
| San Martin de las Piramides | 29,145 |
| Tecamac | 500,585 |
| Temamatla | 13,690 |
| Temascalapa | 41,685 |
| Tenango del Aire | 13,344 |
| Teoloyucan | 69,466 |
| Teotihuacan | 60,992 |
| Tepetlaoxtoc | 33,108 |

| Municipality | Total |
|---|---|
| Tepetlixpa | 21,137 |
| Tepotzotlan | 104,335 |
| Tequixquiac | 39,658 |
| Texcoco | 262,015 |
| Tezoyuca | 46,527 |
| Tlalmanalco | 51,370 |
| Tlalnepantla de Baz | 756,537 |
| Tonanitla | 10,960 |
| Tultepec | 160,943 |
| Tultitlan | 556,493 |
| Valle de Chalco Solidaridad | 419,700 |
| Villa del Carbon | 50,614 |
| Zumpango | 217,166 |

**Figure S1.** Municipalities of Mexico City, State of Mexico and the State of Hidalgo that conform Mexico City Metropolitan Area

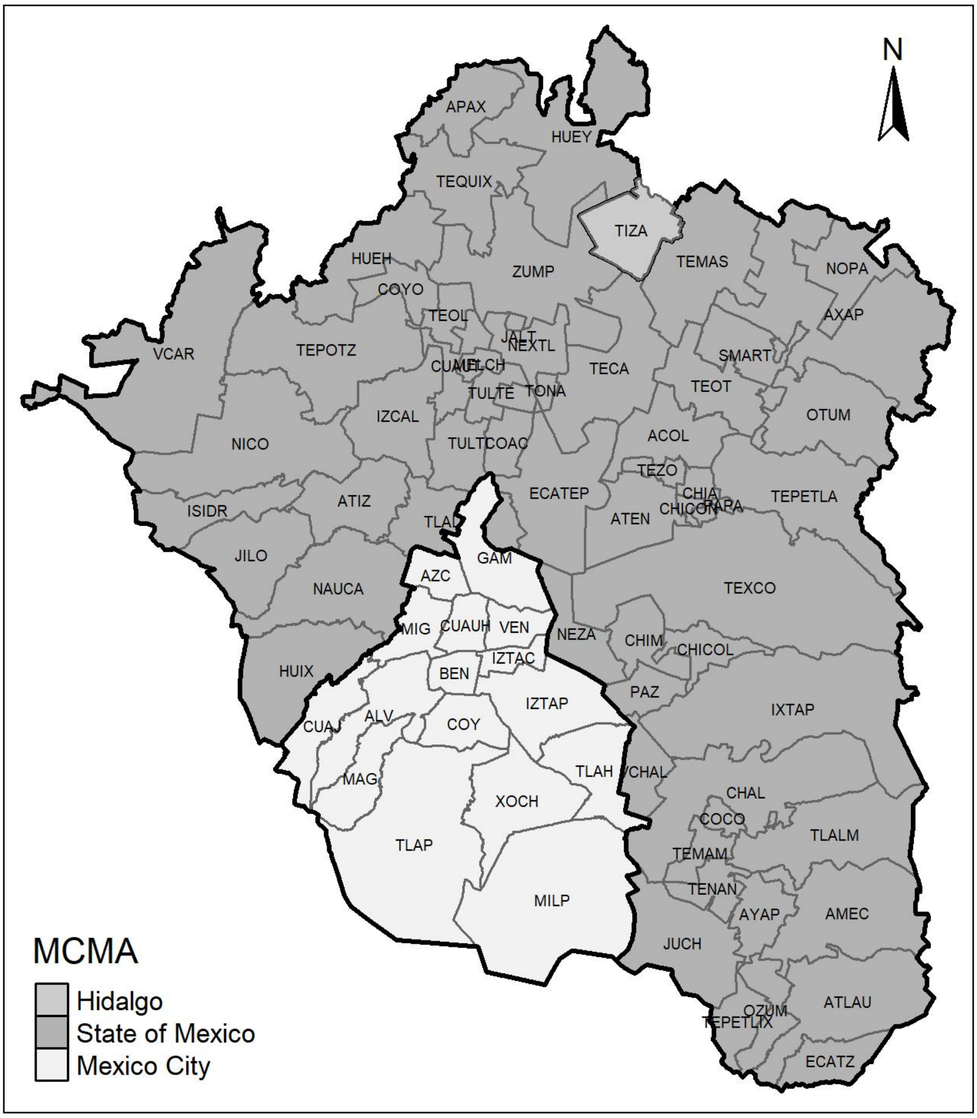

**2. Calibrated posterior distributions**

**Figure S2.** Scatter plot of pairs of calibrated parameters with correlation coefficient and posterior marginal distributions. $\beta$: community transmission rate; $\tau$: household transmission rate; $\eta_1$: effectiveness of NPI on 2020/03/21 - 2020/04/17; $\eta_2$: effectiveness of NPI on 2020/04/17 - 2020/05/23; $\eta_3$: effectiveness of NPI on 2020/05/23 - 2020/08/11; $\eta_4$: effectiveness of NPI on 2020/08/11 - 2020/10/26; $\eta_5$: effectiveness of NPI on 2020/10/26 - 2020/12/07; $\nu_{lb}$: initial detection rate; $\nu_{ub}$: final detection rate; $\nu_{rate}$: rate of change between initial and final detection rate; $\nu_{mid}$: Day at which detection rate is between initial and final values.

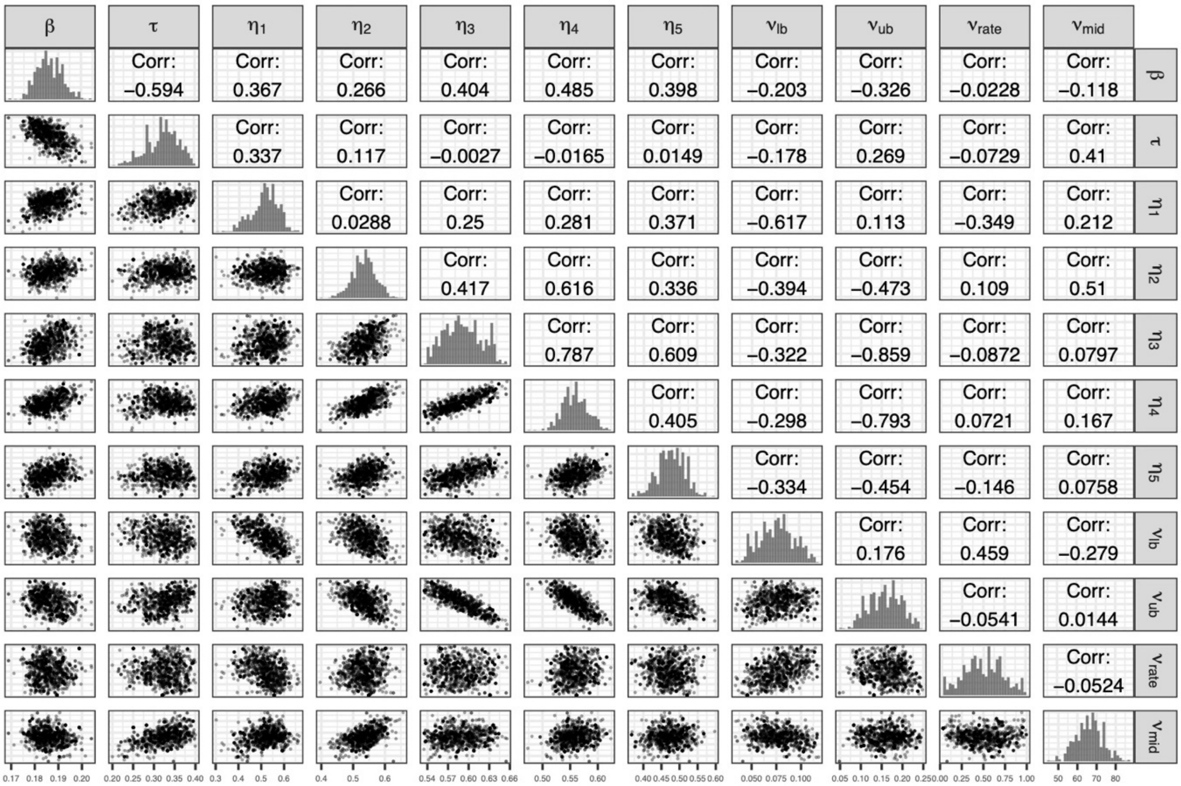

**Figure S3.** Prior and posterior marginal distributions of calibrated parameters. $\beta$: community transmission rate; $\tau$: household transmission rate; $\eta_1$: effectiveness of NPI on 2020/03/21 - 2020/04/17; $\eta_2$: effectiveness of NPI on 2020/04/17 - 2020/05/23; $\eta_3$: effectiveness of NPI on 2020/05/23 - 2020/08/11; $\eta_4$: effectiveness of NPI on 2020/08/11 - 2020/10/26; $\eta_5$: effectiveness of NPI on 2020/10/26 - 2020/12/07; $\nu_{lb}$: initial detection rate; $\nu_{ub}$: final detection rate; $\nu_{rate}$: rate of change between initial and final detection rate; $\nu_{mid}$: Day at which detection rate is between initial and final values.

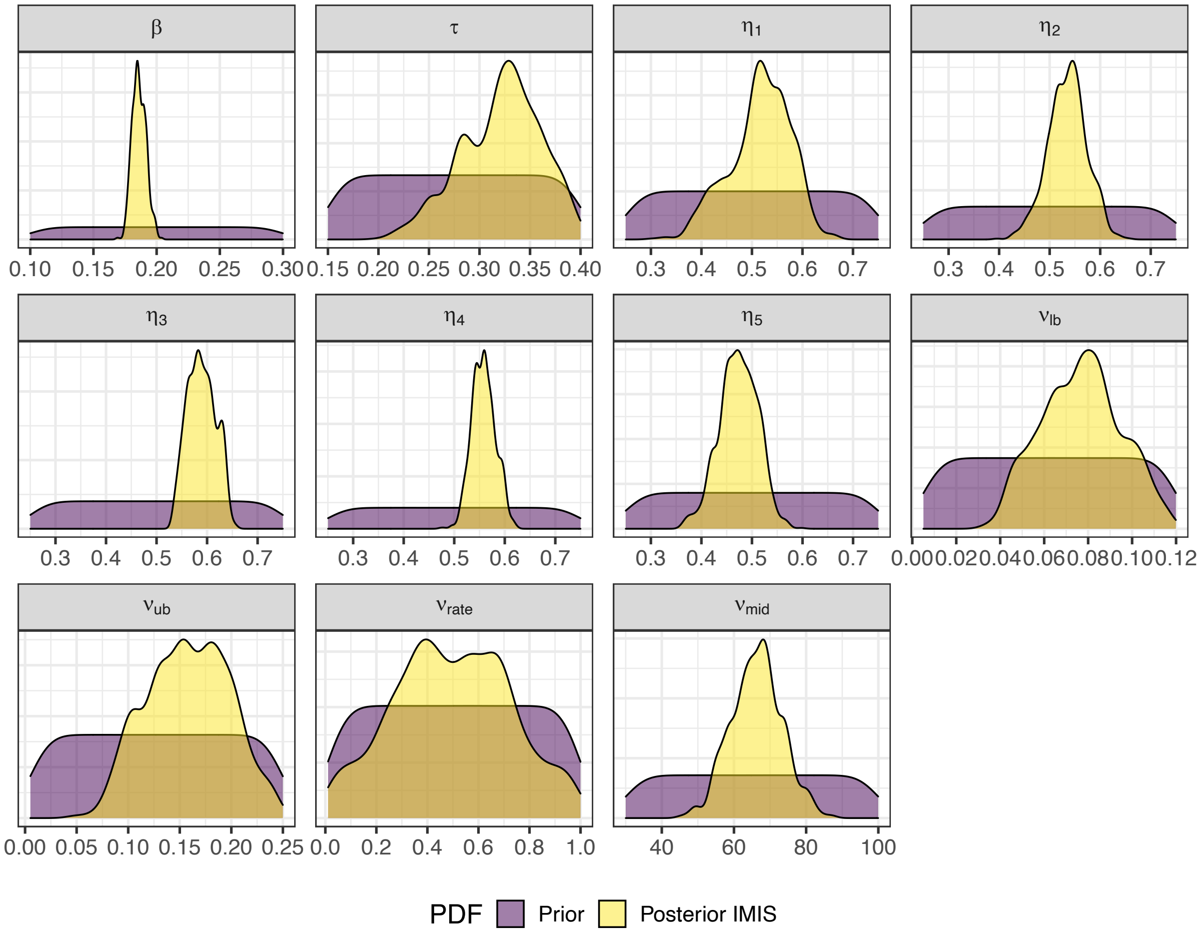

#### 3. Supplementary results

**Figure S4.** Effective reproduction number ($R_e$) for status-quo (A) and by intervention (B) by levels of compliance with social distancing during end-of-year holiday period. Double-dashed vertical line in panel A represents that last day used for calibration. The shaded area shows the 95% posterior model-predictive interval of $R_e$ and colored lines show the posterior model-predicted mean based on 1,000 simulations using samples from posterior distribution.

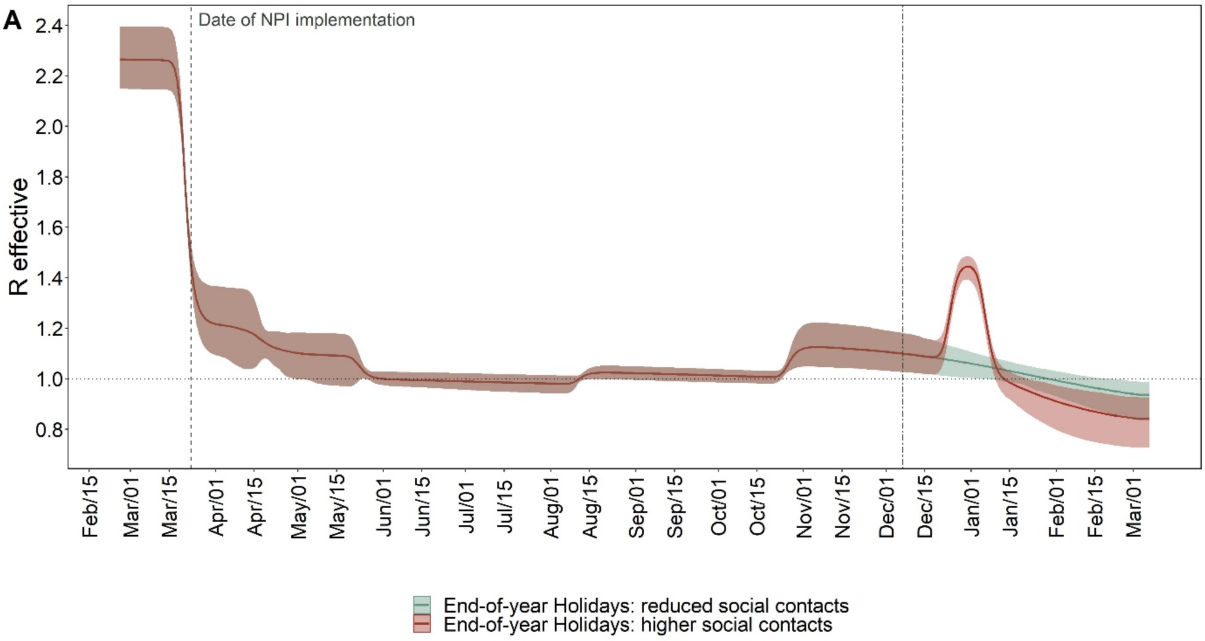


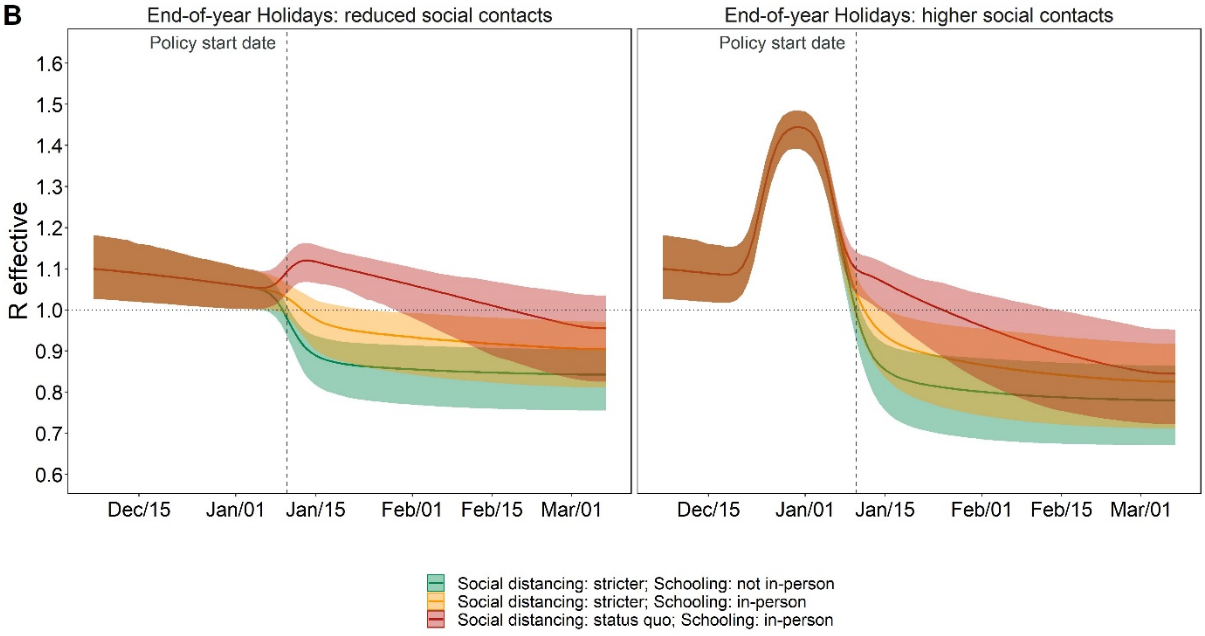

**Figure S5.** Cumulative proportion of population ever been infected under status quo in which there is substantially less compliance with social distancing during the end-of-year holiday period. Double-dashed line indicates last day used for calibration. The shaded area shows the 95% posterior model-predictive interval of the outcomes and the colored line shows the posterior model-predicted mean based on 1,000 simulations using samples from posterior distribution.

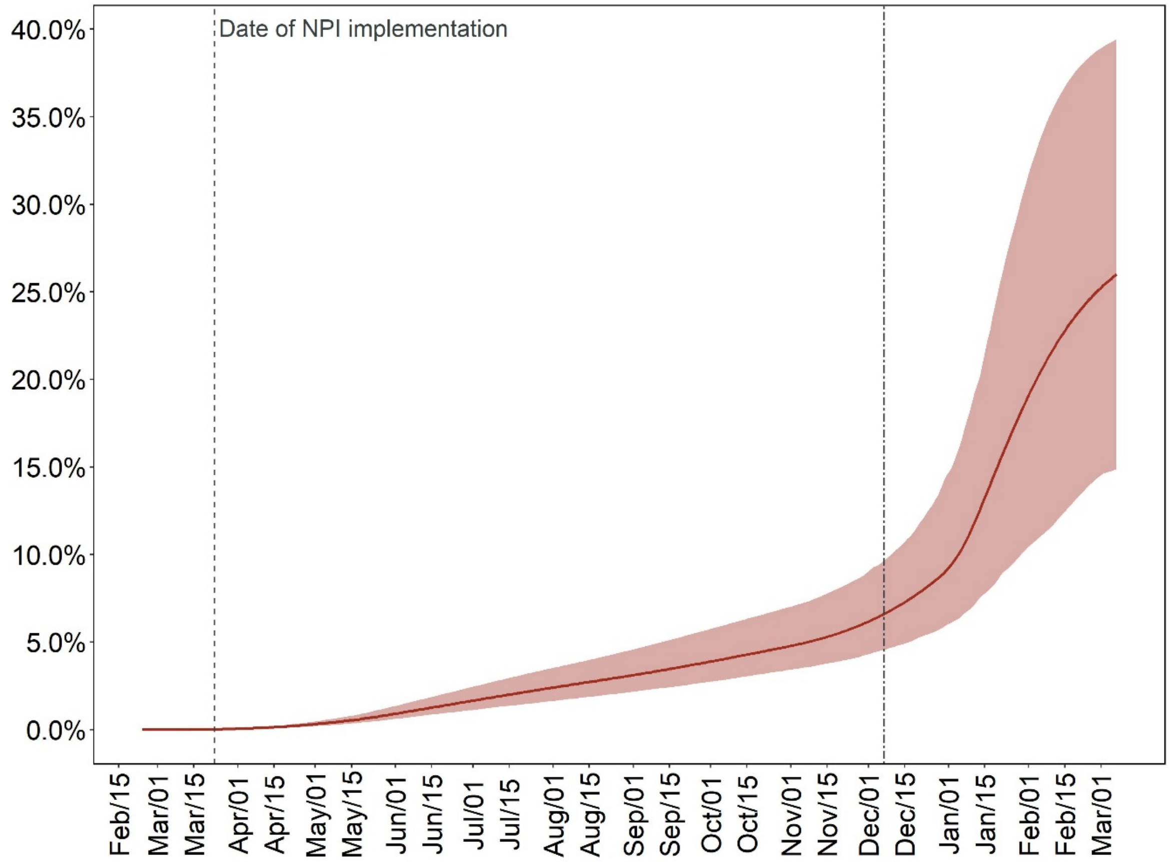

**Figure S6.** Cumulative proportion of infections being detected as cases for the status quo in which there is substantially less compliance with social distancing during the end-of-year holiday period. The model's calibrated case detection rate is time-varying. The shaded area shows the 95% posterior model-predictive interval of the outcomes and the colored line shows the posterior model-predicted mean based on 1,000 simulations using samples from posterior distribution.

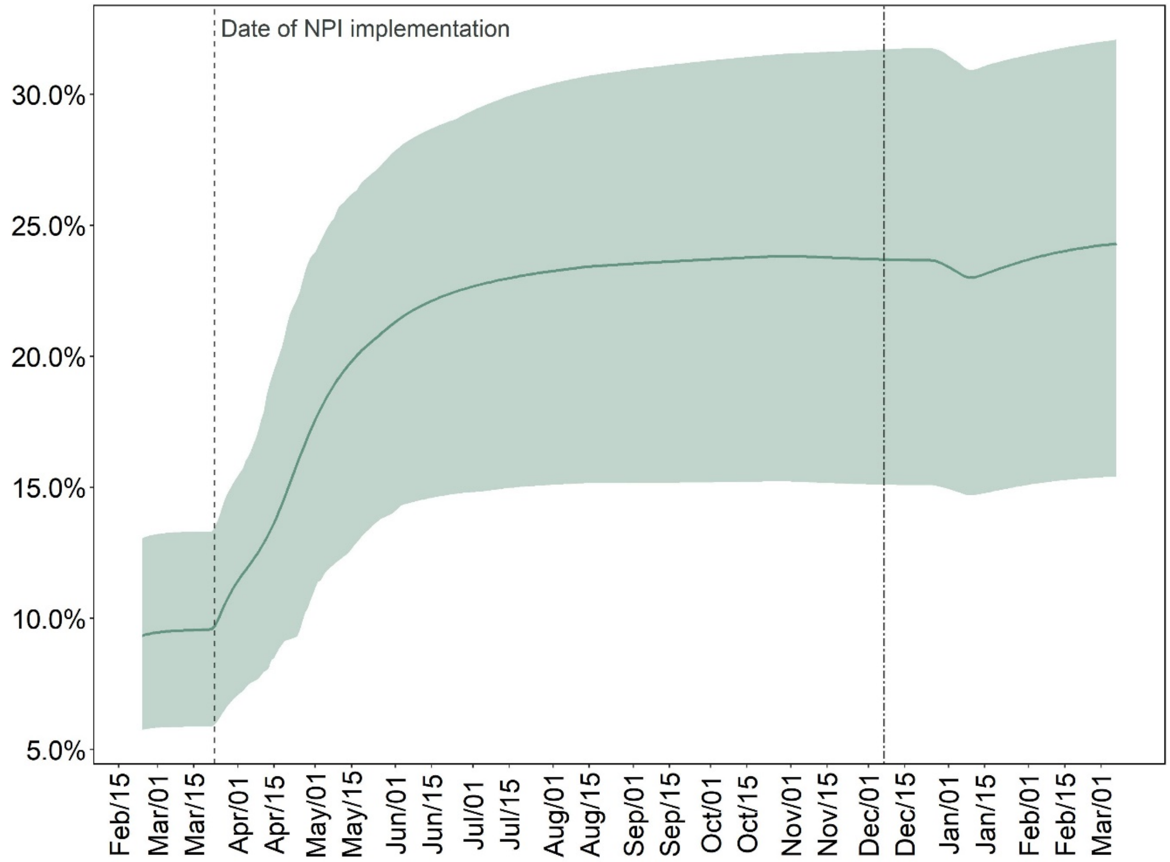

**Table S2.** Estimated mean and 95% prediction interval in parentheses of cumulative cases and deaths by March 07, 2021 under different holiday contact scenario. (A) Assuming reduced social contacts during the end-of-year holiday period; (B) assuming higher social contacts during the end-of-year holiday period.

| **Policy** | **Holiday contact scenario** | |
|---|---|---|
| | **(A)** | **(B)** |
| **Cumulative Covid-19 cases (millions)** | | |
| Social distancing: status quo; Schooling: not in-person | 0·98<br>(0·59 – 1.59) | 1·35<br>(0·79 – 2·00) |
| Social distancing: stricter; Schooling: not in-person | 0·77<br>(0·53 – 1·17) | 1·10<br>(0·70 – 1·59) |
| Social distancing: stricter; Schooling: in-person | 0·87<br>(0·58 – 1·34) | 1·20<br>(0·79 – 1·78) |
| Social distancing: status quo; Schooling: in-person | 1·19<br>(0·66 – 1·92) | 1·61<br>(0·93 – 2·37) |
| **Cumulative Covid-19 deaths (thousands)** | | |
| Social distancing: status quo; Schooling: not in-person | 53<br>(37 – 78) | 68<br>(45 – 94) |
| Social distancing: stricter; Schooling: not in-person | 45<br>(35 – 61) | 57<br>(42 – 78) |
| Social distancing: stricter; Schooling: in-person | 49<br>(37 – 67) | 63<br>(46 – 86) |
| Social distancing: status quo; Schooling: in-person | 61<br>(40 – 89) | 77<br>(50 – 107) |

**Figure S7.** Estimated weekly percent increase in incident and cumulative cases and deaths comparing status-quo in which there is substantially less compliance with social distancing during the end-of-year holiday period vs continuing observed compliance during the holiday period in MCMA. Vertical lines denote the start and end of less compliance with social distancing during the end-of-year holiday period. The error bars show the 95% posterior model-predictive interval of the percentage increases based on 1,000 simulations using samples from posterior distribution.

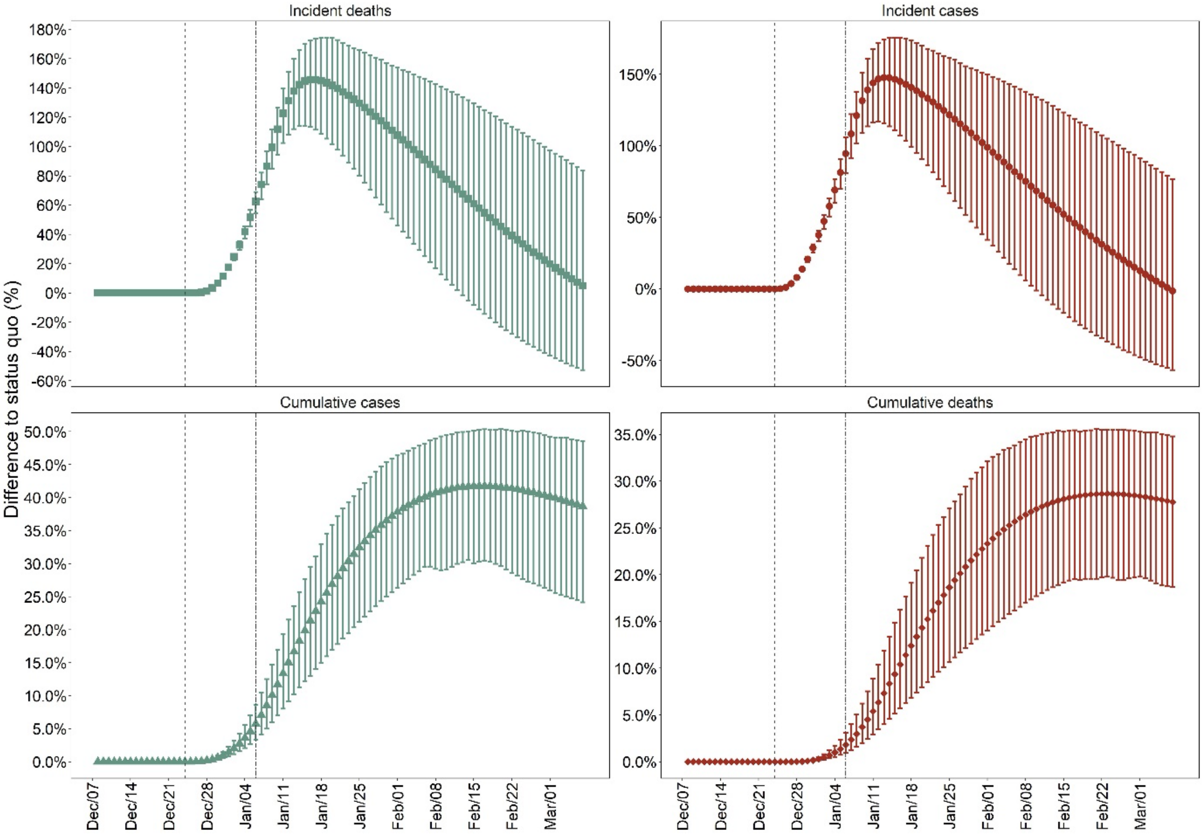

### 4. Stanford-CIDE Coronavirus Simulation Model (SC-COSMO) framework*

*Additional contributors to the specific design and implementation of the SC-COSMO model described below are listed in alphabetical order and include: Anneke Claypool, Michael Fairley, Valeria Gracia, Natalia Kunst, Andrea Luviano, Yadira Peralta, Marissa Reitsma.

†All Members of the SC-COSMO Modeling Consortium provided support, input and/or helpful comments on our work generally and are listed in alphabetical order here: Fernando Alarid-Escudero, Jason Andrews, Jose Manuel Cardona Arias, Liz Chin, Anneke Claypool, Hugo Berumen Covarrubias, Ally Daniels, Mariana Fernandez, Hannah Fung, Zulema Garibo, Jeremy Goldhaber-Fiebert, Valeria Gracia, Alex Holsinger, Erin Holsinger, Radhika Jain, Neesha Joseph, Natalia Kunst, Elizabeth Long, Andrea Luviano, Regina Isabel Medina Rosales, Marcela Pomar Ojeda, Yadira Peralta, Lea Prince, Marissa Reitsma, Neil Rens, Tess Ryckman, Joshua Salomon, David Studdert, Hirvin Azael Diaz Zepeda.

#### 1 Model description

The epidemiology of COVID-19 in the absence of treatment or vaccination can be described as a multi-compartment susceptible-exposed-infected-recovered-susceptible (MC-SEIR) model with demography.[1] In such a model, the exposed ($E$) compartments represent individuals that are infected but who are not yet infectious, and the infectious ($I$) compartments represent individuals that are both infected and infectious (i.e., can infect susceptibles ($S$) if a contact occurs).[1] Notably, by using multiple levels for the $E$ and $I$ compartments along with allowing for differential rates of symptom onset and of detection (described in sections below), the model structure can also capture the possibility of asymptomatic infectiousness as well as infectiousness varying over the course of infection. Figure S8 depicts the generalized structure of the non-age-stratified MC-SEIR compartmental model (age structure described below). Each of these compartments represents part of the population characterized by their COVID-19 status as a function of time.

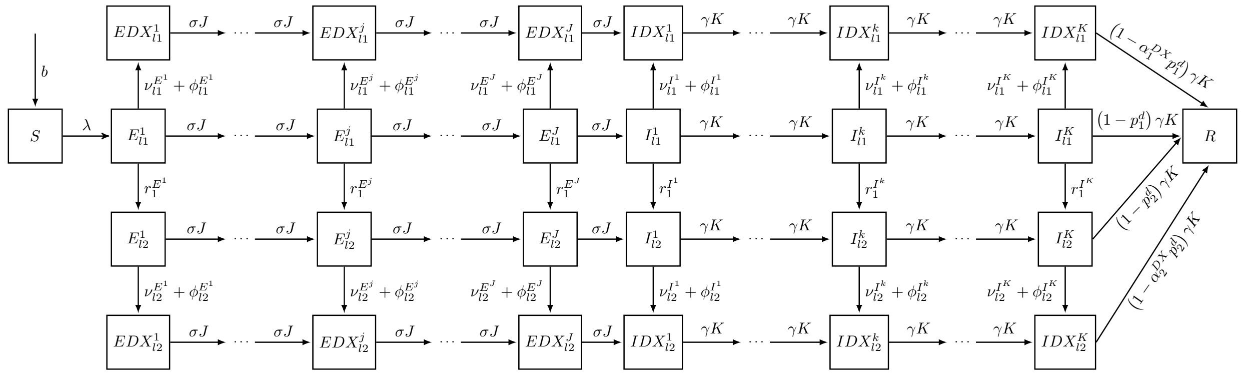


*Note: Background mortality rate ($\mu$) from all model compartments (not shown)

Figure S8: Diagram of the SC-COSMO model.

Figure S8 includes a number of important processes and for simplicity elides others that are included in the model and that we will describe subsequently. In the figure, we see that people are born into the susceptible compartment $S$ at a rate $b$. Not shown explicitly, all people face age-specific background mortality rates ($\mu$) from all compartments. People can become infected at a rate $\lambda$ and enter the exposed compartment $E$, progressing to becoming infectious $I$ before recovering and entering $R$. Those who are infected face an excess risk of death from their COVID-19 infections which may be reduced by proper and timely supportive medical care ($p^d$ and $\alpha^{DX}$, respectively). The figure shows that the exposed and infectious compartments are stratified by severity level ($l = 1, \ldots, L$) and by whether they have been diagnosed ($DX$). We ensure realistic distributions of times in $E$ and $I$ by using a multi-compartment structure; specifically, we have multiple $E$ and $I$ compartments, and rates of progression ($\sigma$) from $E$ to $I$ and ($\gamma$) from $I$ to $R$ that are multiplied by the number of each type of compartments ($J$ and $K$, respectively).[1]

For epidemics like COVID-19, both age-structure and household-structure are important to consider to appropriately capture its dynamics.[2] We next describe how these are incorporated into the model.

We begin with age-structure. Because it is important to model the age-dependent dynamics of COVID-19 contacts, transmission, and case severity, we expanded the MC-SEIR model to include a realistic age structure (RAS) and heterogeneous, venue-specific age-structured mixing outside of the household (we describe household mixing later). We divided the population into $N$ age groups where each $a$-th age group has its own set of $S_a$, $E^j_{l,a}$, $EDX^j_{l,a}$, $I^k_{l,a}$, $IDX^k_{l,a}$, and $R_a$ compartments for $a = 1, \ldots, N$. The total size of the population at time $t$ is $Pop$ which is the sum across compartments, age groups, and all other stratifications noted above. Importantly, we term the components of the model described up until now the "community submodel" (i.e., the non-household components) to differentiate them from the "household submodel" which we describe next.

We provide an initial description of the household submodel and how it interacts with the community submodel. The household submodel acknowledges that people are generally embedded within households. Such embeddings are important to consider particularly because interventions have focused on shelter-in-place where people continue to be exposed to contacts and transmission within their households even as their exposures outside of their households have been reduced.

It is challenging to construct compartmental models that properly embed within-household and community transmission. Approaches for doing this have been described previously for simple SIR models (e.g.,[3]). We extend the prior approach in,[3] combining it with our community submodel (the RAS MC-SEIR model) for COVID-19 to produce an overall model of the population.

To describe overall transmission dynamics in the population, we first describe the key elements of the household submodel, then the details of transmission in the RAS MC-SEIR submodel and how the household submodel's transmission is integrated into it, and finally provide full details about the household submodel. The key components of the household submodel are: 1) once a given household's members are all infected and/or recovered, no further transmission occurs within that household (unless there are births into the household); and 2) if households are not completely isolated from one another such that community transmission is still occurring, then the within-household force of infection (related to the household Secondary Attack Rate, e.g.,[4]) can drive additional community transmission. This occurs through chains of transmission where one household member infects another and then, either member also transmits to individuals in the community who are outside of the household. Through these key features, the household submodel generates a component of the overall force of infection which we term $\lambda_{HH}$. The details of this force of infection are provided with the description of the household submodel below in equation (11).

#### 1.1 Community submodel with transmission from household force of infection

The community submodel (the RAS MC-SEIR model) is described by a system of $[(2 + 2L(J + K))N]$ ordinary differential equations (ODEs):

$$
\begin{aligned}
\frac{dS_1}{dt} &= bPop - (\lambda_1 + \mu_1)\,S_1 - \lambda_{1,HH}Pop_1\\
\frac{dS_a}{dt} &= -(\lambda_a + \mu_a)\,S_a - \lambda_{a,HH}Pop_a \text{ for } a = 2,\dots,N\\
\frac{dE^1_{1,a}}{dt} &= \lambda_a S_a - \left(r^{E^1}_{1,a} + \sigma J + \nu^{E^1}_l + \phi^{E^1}_l + \mu_a\right) E^1_{l,a} + \lambda_{a,HH}Pop_a\\
\frac{dE^j_{l,a}}{dt} &= \sigma J E^{j-1}_{l,a} + r^{E^j}_{(l-1),a}E^j_{(l-1),a} - \left(r^{E^j}_{l,a} + \sigma J + \nu^{E^j}_l + \phi^{E^j}_l + \mu_a\right) E^j_{l,a} \text{ for } j = 2,\dots,J;\ l = 2,\dots,(L-1)\\
\frac{dE^j_{L,a}}{dt} &= \sigma J E^{j-1}_{L,a} + r^{E^j}_{(L-1),a}E^j_{(L-1),a} - \left(\sigma J + \nu^{E^j}_L + \phi^{E^j}_L + \mu_a\right) E^j_{L,a} \text{ for } j = 2,\dots,J\\
\frac{dEDX^1_{l,a}}{dt} &= \left(\nu^{E^1}_l + \phi^{E^1}_l\right) E^1_{l,a} - (\sigma J + \mu_a)\,EDX^1_{l,a}\\
\frac{dEDX^j_{l,a}}{dt} &= \sigma J EDX^{j-1}_{l,a} + \left(\nu^{E^j}_l + \phi^{E^j}_l\right) E^j_{l,a} - (\sigma J + \mu_a)\,EDX^j_{l,a}, \text{ for } j = 2,\dots,J;\ l = 1,\dots,L\\
\frac{dI^1_{1,a}}{dt} &= \sigma J E^J_{l,a} - \left(r^{I^1}_{1,a} + \gamma K + \nu^{I^1}_l + \phi^{I^1}_l + \mu_a\right) I^1_{l,a}\\
\frac{dI^k_{l,a}}{dt} &= \gamma K I^{k-1}_{l,a} + r^{I^k}_{(l-1),a}I^k_{(l-1),a} - \left(r^{I^k}_{l,a} + \gamma K + \nu^{I^k}_l + \phi^{I^k}_l + \mu_a\right) I^k_{l,a} \quad \text{for } k = 2,\dots,K;\ l = 2,\dots,(L-1)\\
\frac{dI^k_{L,a}}{dt} &= \gamma K I^{k-1}_{L,a} + r^{I^k}_{(L-1),a}I^k_{(L-1),a} - \left(\gamma K + \nu^{I^k}_L + \phi^{I^k}_L + \mu_a\right) I^k_{L,a} \text{ for } k = 2,\dots,K\\
\frac{dIDX^1_{l,a}}{dt} &= \sigma J EDX^J_{l,a} + \left(\nu^{I^1}_l + \phi^{I^1}_l\right) I^1_{l,a} - (\gamma K + \mu_a)\,IDX^1_{l,a}\\
\frac{dIDX^k_{l,a}}{dt} &= \left(\nu^{I^k}_l + \phi^{I^k}_l\right) I^k_{l,a} - (\gamma K + \mu_a)\,IDX^k_{l,a}, \text{ for } k = 2,\dots,K;\ l = 1,\dots,L\\
\frac{dR_a}{dt} &= \sum_{l=1}^{L}\left[\left(1 - p^d_{l,a}\right)\gamma K\left(I^K_{l,a}\right) + \left(1 - \alpha^{DX}_l p^d_{l,a}\right)\gamma K\left(IDX^K_{l,a}\right)\right] - \mu_a R_a,
\end{aligned}
\tag{1}
$$

where $b$ is the birth rate into the youngest age class $a = 1$; $\sigma$ is the rate at which exposed individuals in class $E^j_{l,a}$ progress to class $E^{j+1}_{l,a}$ and also from $EDX^j_{l,a}$ to class $EDX^{j+1}_{l,a}$ for $j = 1,\dots,(J-1)$ and from the exposed class $E^J_{l,a}$ to the infected class $I^1_{l,a}$ and from $EDX^J_{l,a}$ to $IDX^1_{l,a}$; $r^{E^j}_{l,a}$ is the rate of developing a more severe infection for individuals moving from $E^j_{l,a}$ to $E^j_{(l+1),a}$ for each of the severity classes $l = 1,\dots,(L-1)$; $\nu^{E^j}_l$ is the rate of detection due to symptoms from which $E^j_{l,a}$ go to $EDX^j_{l,a}$ for $j = 1,\dots,J$; $\phi^{E^j}_l$ is the rate of detection due to screening from which $E^j_{l,a}$ go to $EDX^j_{l,a}$ for $j = 1,\dots,J$; $\gamma$ is the rate at which infectious individuals in class $I^k_{l,a}$ progress to class $I^{k+1}_{l,a}$ and also from $IDX^k_{l,a}$ progress to class $IDX^{k+1}_{l,a}$ for $k = 1,\dots,(K-1)$ and it is also the recovery rate from the infectious classes $I^K_{l,a}$ and $IDX^K_{l,a}$ to the recovered class $R_a$; $r^{I^k}_{l,a}$ is the rate of developing a more severe infection for individuals moving from $I^k_{l,a}$ to $I^k_{(l+1),a}$ for each of the severity classes $l = 1,\dots,(L-1)$; $\nu^{I^k}_l$ is the rate of detection due to symptoms from which $I^k_{l,a}$ go to $IDX^k_{l,a}$ for $k = 1,\dots,K$; $\phi^{I^k}_l$ is the rate of detection due to screening from which $I^k_{l,a}$ go to $IDX^k_{l,a}$ for $k = 1,\dots,K$; $p^d_{l,a}$ is the proportion of infectious individuals in class $I^K_{l,a}$ that die from COVID-19; $\alpha^{DX}_l$ is a reduction in the proportion who die from COVID-19 due to detection and appropriate healthcare in severity class $l$; and $\mu_a$ represents the age-specific background mortality experienced from all compartments and stratifications.

Given its main intended uses, the current model does not include immigration inflows into the population nor does it include population aging. Its equations would require modification to consider such scenarios.

Table 1: Description of variables, subscripts and superscripts

| Symbol | Description |
|---|---|
| Subscripts | |
| $a$ | Age group $\{1,\ldots,N\}$ |
| $l$ | Severity levels $\{1,\ldots,L\}$ |
| Superscripts | |
| $j$ | Number of exposed compartment $\{1,\ldots,J\}$ |
| $k$ | Number of infectious compartment $\{1,\ldots,K\}$ |
| Variables | |
| $\lambda_a$ | Force of infection at age group $a$ overall |
| $\lambda_{a,HH}$ | Force of infection at age group $a$ from household transmission |
| $S_a$ | Susceptible in age group $a$ |
| $E_a^j$ | $j$-th exposed/infected in age group $a$; for $j=1,\ldots,J$ |
| $EDX_{l,a}^j$ | $j$-th detected exposed/infected in age group $a$; for $j=1,\ldots,J$ and $l=1,\ldots,L$ |
| $I_{l,a}^k$ | $k$-th infectious in age group $a$; for $k=1,\ldots,K$ and $l=1,\ldots,L$ |
| $IDX_{l,a}^k$ | $k$-th detected infectious in age group $a$; for $k=1,\ldots,K$ and $l=1,\ldots,L$ |
| $R_a$ | Recovered in age group $a$ |

##### 1.1.1 Force of infection

The force of infection (FOI), $\lambda$, is the key quantity governing the transmission of infection within a given population, defined as the instantaneous per capita rate at which susceptibles acquire infection. FOI reflects both the degree of contact between susceptibles and infectious individuals and the transmissibility of the pathogen per contact. Because contacts can happen in a variety of different venues $(1,...,V)$ that may be differentially reduced under particular intervention scenarios (e.g., school closures reduce school contacts), we actually construct $\lambda$ as $\sum_{v=1}^{V}\lambda^v$ for all non-household venues. The model also incorporates a force of infection from household transmission, $\lambda_{HH}$, which we define separately from the other components of the FOI in the description of the household submodel below in equation (11).

The FOI $\lambda_a^v$ (for the non-household venues) represents the venue-specific rate of disease transmission from infectious people in all age groups to susceptibles in age group $a$,[5] and likewise, we define the age-group specific FOI for household transmission $\lambda_{a,HH}$. Overall, the non-household $\lambda_a^v$ is defined for the COVID-19 RAS MC-SEIR model from a particular venue $v$ as

$$\lambda_a^v = \sum_{k=1}^{K}\left[\sum_{a'=1}^{N}\beta_a^k W_{a,a'}^v\left(\sum_{l=1}^{L}\frac{I_{l,a'}^k}{Pop}\right) + \sum_{a'=1}^{N} f\beta_a^k W_{a,a'}^v\left(\sum_{l=1}^{L}\frac{IDX_{l,a'}^k}{Pop}\right)\right]; \quad a=1,\ldots,N, \tag{2}$$

where the transmission rate, $\beta_a^k$, describes the probability that an infected individual of age $a'$ who is $k$ days into his infectious period will infect a susceptible of age $a$ per unit of time and $W_{a,a'}^v$ is the $\{a,a'\}$ entry of the venue-specific Who-Acquired-Infection-From-Whom (WAIFW) matrix, $W^v$, and $f\in[0,1]$ is a reduction factor in transmission from infectious individuals that are diagnosed (due for example to quarantine and isolation). As shown in equation 2 by the fact that we are summing over infection severity levels $l$ and that we do not have separate $\beta$ parameters by severity level, the current model makes no assumption about severity level-specific differential transmissibility other than the indirect effect that more severe infections are more likely to be diagnosed and hence may transmit less frequently post diagnosis. In fact, the current model assumes that $\beta$ is a single constant value.

Each $W^v$ has $N^2$ elements, representing mixing between each pair of age groups in the model at that venue. The venue-specific FOI in equation (2) for $\lambda^v$ is therefore a system of $N$ equations that can be represented in matrix form.

$$\lambda^v = \beta W \frac{I}{Pop} + f \beta W \frac{IDX}{Pop}$$

$$\begin{bmatrix} \lambda_1^v \\ \lambda_2^v \\ \vdots \\ \lambda_N^v \end{bmatrix} = \beta \begin{bmatrix} W_{1,1}^v & W_{1,2}^v & \cdots & W_{1,N}^v \\ W_{2,1}^v & W_{2,2}^v & \cdots & W_{2,N}^v \\ \vdots & \vdots & \ddots & \vdots \\ W_{N,1}^v & W_{N,2}^v & \cdots & W_{N,N}^v \end{bmatrix} \begin{bmatrix} \sum_{k=1}^K I_{l,1}^k / Pop & \sum_{k=1}^K IDX_{l,1}^k / Pop \\ \sum_{k=1}^K I_{l,2}^k / Pop & \sum_{k=1}^K IDX_{l,2}^k / Pop \\ \vdots & \vdots \\ \sum_{k=1}^K I_{l,N}^k / Pop & \sum_{k=1}^K IDX_{l,N}^k / Pop \end{bmatrix} \begin{bmatrix} 1 \\ f \end{bmatrix} \quad (3)$$

#### 1.2 Household submodel

As with,[3] the household submodel tracks the proportion of households whose members are in various disease states of COVID-19's natural history. For example, of all households in a given population at some time during the simulation, 5% of 3-person households might have 1 member susceptible and 2 members recovered ($HH_{(S=1,...,R=2)}$). More generally, we denote the proportion of households whose members are in any combination of natural history states (i.e., counts of members in each state which we abbreviate $sc$ for state counts) as $HH_{sc}$ where $1 = \sum_{sc} HH_{sc}$.

The number of distinct proportions ($HH_{sc}$) that represent households with different state counts grows rapidly with both household size ($hhsize$) and with the number of natural history states ($states$). In fact, the equation for the number of household proportions (and hence differential equations) with a fixed household size is:

$$ODEs = \frac{(hhsize + states - 1)!}{(hhsize!(states - 1)!)}. \quad (4)$$

To keep the number of household types, and hence ODEs, manageable, we make a number of simplifying assumptions. First, we assume that all households are the same size as the average household for a given location, rounding the average household size to the nearest whole integer (e.g., $hhsize = 3$ for counties in California and likewise for Mexico City Metropolitan Area, Mexico). Second, we assume that the age composition, distribution of infection severity, and fraction of infections that are detected are the same across households; therefore, while the household submodel retains the multi-compartment structure for exposed $E$ and infectious $I$ states, it does not explicitly stratify by age of household members, nor does it explicitly differentiate between undetected and detected ($DX$) infections or severity levels of infection.

The household submodel's initial state is computed in a manner that corresponds with the community submodel's initial state. For a given size of the total population at the start of the community model $Pop_{t=0}$ under the assumption of all households being size $hhsize = hhsize_{avg}$, the number of households is $N_{households} = Pop_{t=0}/hhsize_{avg}$. If there is one person in the $E_1$ state in the entire population, then the fraction of households that have an infected member is $1/N_{households}$ and the remainder are households with all susceptibles: $(N_{households} - 1)/N_{households}$. For other starting conditions (i.e., more than one exposed, infectious, or recovered individual) this initialization generalizes easily under the assumption (which we make) that the initial few infections are not correlated within household (i.e., if there are 3 infections they would be in 3 separate households).

The household submodel's dynamics include: progression, recovery, within-household and community-household transmission, births, and deaths. Modeling many of these dynamics in the household submodel is somewhat more complicated than in the community submodel because the household submodel tracks the fraction of households in a set of discrete states characterized by counts of members in each natural history state and has multiple exposed and infectious compartments relevant for progression and recovery.

The intuition of how the household submodel handles progression and recovery is given in the following set of examples. In a simplified example ignoring the multi-compartment nature of the exposed and infectious states and considering only progression, if there are 4 household members (1 susceptible, 3 exposed, 0 infectious) at a given time, then it is possible that 0, 1, 2, or all 3 of the exposed members will progress to infectious on a given day. Hence, the possible states that this household could go to include (1 susceptible, 3 exposed, 0 infectious), (1 susceptible, 2 exposed, 1 infectious), (1 susceptible, 1 exposed, 2 infectious), or (1 susceptible, 0 exposed, 3 infectious). In the example, the frequency of households moving to each of the states follows a binomial distribution with the probability related to the rate of progression ($\sigma$). In a similar example considering multiple exposed and infectious compartments in the MC-SEIR model, the binomial distribution's probability is then related to $\sigma J$; and if there are household members in several of the multi-compartment exposed states, the general form of these resulting frequencies of household states follows a convolution of binomial distributions. This is likewise the case for recovery where, in the multi-compartment model, the frequencies of resulting household states also follow a convolution of binomial distributions with probability related to $\gamma K$.

In the examples for progression and recovery above for models of counts of household members where $E$ and/or $I$ are multi-compartment, resulting counts of household members in states follow a convolution of binomial distributions. The details of this calculation in a general form are given here. Consider $C$ individuals (i.e., the members of a household) each in a Markov chain with states $X_t^c \in \{1, \ldots, M\}$ for $c \in \{1, \ldots, C\}$. The $M$ states in our case are those in the MC-SEIR model. The Markov chain has the following transition probabilities (where for simplicity we have set the probability of flow from R to S equal to 0):

$$P(X_{t+1}^c = X_t^c + 1 \mid X_t^c) = p, \quad X_t = 1, \ldots, M-1 \tag{5}$$

$$P(X_{t+1}^c = X_t^c \mid X_t^c) = 1 - p, \quad X_t = 1, \ldots, M-1 \tag{6}$$

$$P(X_{t+1}^c = X_t^c \mid X_t^c) = 1, \quad X_t = M \tag{7}$$

In other words, for all states except the last, with probability $p$, each individual progresses from $X_t$ to $X_{t+1}$ (e.g., $E_2$ to $E_3$ or from $E_3$ to $I_1$) and with probability $1-p$ the individual stays in the same state. Individuals remain with certainty in the last ($M^{th}$) state after progressing to it.

Having considered each individual, we now consider counts of household members. Let $Y_t^m$ be the number of individuals in state $m$ at time $t$. We consider a new Markov chain with state $(Y_t^1, \ldots, Y_t^M)$. The transition probabilities can be calculated as follows. Given a transition from state $(Y_t^1, \ldots, Y_t^M)$ to state $(Y_{t+1}^1, \ldots, Y_{t+1}^M)$:

1. For each of the $M-1$ transition arcs in the underlying Markov chain, find the number of individuals transitioning from state $m$ to $m+1$, denoted $\Delta_t(m, m+1)$ for $m < M$.

2. The probability of the transition is then a (convolution of) binomial distribution(s):

$$\prod_{m=1}^{M-1} \binom{Y_t^m}{\Delta_t(m, m+1)} p^{\Delta_t(m,m+1)} (1-p)^{Y_t^m - \Delta_t(m,m+1)} \tag{8}$$

To find the value of $\Delta_t(m, m+1)$, use the following backwards recursion:

$$\Delta_t(M-1, M) = Y_{t+1}^M - Y_t^M \tag{9}$$

$$\Delta_t(m-1, m) = Y_{t+1}^m + \Delta_t(m, m+1) - Y_t^m \tag{10}$$

In other words, for each source state (e.g., $E_2$) which at time $t$ might have $h$ household members in it, 0, 1, ..., $h$ members may progress to $E_3$ at time $t+1$ with the rest remaining in $E_2$. The counts of progressors are binomial distributed. However, if there are also some household members in $E_3$ at time $t$ then the count of people in $E_3$ at time $t+1$ is more complicated because it depends on the count of the progressors from the first example as well as the count of the non-progressors among those in $E_3$. Hence, we arrive at a convolution of binomial distributions as the general description provided in equation (8) with a simple binomial distribution for cases where there are individuals in one source state and none in the destination state at time $t$.

There are both within-household and community-household transmission routes in the submodel. Within-household transmission involves infectious household members infecting susceptible household members. Community-household transmission involves infectious individuals in the community (people who are not household members) infecting susceptible household members. Within-household transmission is related to three components: a) the current number of infectious household members; b) the rate of contact between household members; c) the probability of within-household transmission given household contacts ($\tau$). The number of infectious individuals in the household is given directly by the household compartment being considered. The number of daily household contacts is computed from the household mixing matrix for the given jurisdiction that we estimate as described below. Finally, note that because the intensity of household contacts may differ from contacts in the community, the probability of transmission conditional on household contacts differs from the probability of transmission given community contacts ($\beta$) that we described above.

Like the community submodel, the household submodel also includes birth and deaths but in a manner that is more simplified. First, since the household submodel considers proportions of households, it assumes that births equal deaths since we want the sum of the proportions to equal 1 at all times (i.e., the number of households may increase even as the proportions remain the same). Second, the household submodel assumes that the fraction of all deaths that are due to COVID-19 is relatively small and hence exposes households to an average background mortality rate (consistent with the jurisdiction that the community model is representing). Hence, there is an outflow from all household submodel compartments at this death rate proportional to the fraction of all households they represent, which determines an inflow of births that is spread proportionally across household compartments representing only those with at least one susceptible member (i.e., newly born individuals are assumed to be born susceptible).

The household submodel must make certain approximations because it does not explicitly stratify by age structure nor does it do so by diagnosis status of infectious individuals. This has several implications. The first is that contact rates with household members and also with community members are age-weighted averages of sums of contacts across ages in the corresponding venue-specific WAIFW matrices that are described above yielding $contacts_{HH}$. The second is that the community force of infection in the household submodel comprises age-weighted averages of forces of infection from infectious and detected infectious individuals in the community submodel. The third is that just as $\beta$ is scaled by $f$ in the community submodel for individuals who are infectious and detected, so too $\tau$ is scaled by the same constant for within-household transmission yielding $\tau'$. However for scaling $\tau$ to produce $\tau'$, the fraction of household infectious contacts to which this scaling factor applies is assumed to be the same as the fraction of prevalent infectious individuals who are currently detected in the community submodel.

With this description of the household submodel, we can now define the household force of infection, $\lambda_{HH}$, which connects the dynamics of the household submodel back to the community submodel. $\lambda_{HH}$ depends on the number of within-household contacts between susceptible and infectious members and the probability of transmission given a household contact ($\tau'$). Within each household, we define the rate of new infections:

$$infection_{HH} = \tau' * contacts_{HH} * \sum_{sc} HH_{sc} \left( \frac{infectious_{HH,sc} * susceptible_{HH,sc}}{hhsize} \right), \tag{11}$$

which is the weighted average of within-household transmission (higher where there are more infectious and susceptible individuals simultaneously present in the household) where the weight is the fraction of households who have these counts of infectious and susceptible members. The infections generated by household transmission, $infection_{HH}$, are not age stratified nor are they scaled to the overall population size so we multiply the rate by the size of the population and then spread the new infections flowing from $S_a$ to $E_a$ based upon the proportion of the overall susceptible population in each age group $a$ at time $t$. This produces $\lambda_{a,HH}$ which we include in the community submodel above.

We presented further details of this approach at the 2020 Society for Medical Decision Making Annual Meeting.[6]

##### 1.2.1 Epidemiologic parameters

The epidemiology of COVID-19 is being elucidated at a rapid rate. Key epidemiological parameters include the latent period (i.e., time spent infected but not yet infectious); the incubation period (i.e., time spent infected but without symptoms); and the infectious period (time spent infectious prior to recovery). Data from[7–9] suggest that there are periods of time where some individuals are infectious but asymptomatic and also where some individuals are symptomatic but not yet infectious.

We use the Exposed (E) compartments and Infectious (I) compartments to capture the latent and infectious periods, estimating the distribution of their duration based primarily on.[8] Specifically, we obtained the data and code from the publications to regenerate the empirical distribution of outcomes in their sample. Then, we fit gamma distributions for the distribution of durations of the latent and infectious periods respectively (latent shape and rate parameters (9.00, 3.00); infectious shape and rate parameters (2.18, 0.70)). These yield mean durations of latency and infectiousness of 3 days ($1/\sigma$) and 3.12 days ($1/\gamma$) respectively. For the latent period, 95 percent of people have durations between 1.5 and 4.9 days. For the infectious period, 95 percent of people have durations between 0.6 and 7.0 days.

We need to determine the numbers of E and I compartments for our MC-SEIR model. This is important because the distributions of duration in a compartment are not exponential (what will result if there are only single E and I compartments respectively) but rather to be realistic and consistent with the empirical data by having multiple compartments they will be gamma distributed.[1] The number of compartments for E and for I must be positive integers and should in general be less than or equal to the average duration. We sample from the gamma distribution that is parametrized in terms of number of compartments and average duration from,[1] selecting the number of compartments whose distribution is as close to those fit to the data from[8] in the previous step (i.e., we minimize the sum of square errors across the support of the distributions). We find that the best fit is achieved with 3 E compartments and 2 I compartments.

To connect symptoms to our exposed compartments (E) and infectious compartments (I), we use the idea of severity classes $l$ that index the E and I compartments to denote asymptomatic ($l = 1$) and symptomatic ($l = 2$) individuals. Similar to how we estimate durations of latency and infectiousness, we estimate the incubation period as a gamma distribution, fitting to the data from[7] (incubation shape and rate parameters (2.97, 0.59)) and also considering.[9] This yields a mean duration of incubation of 3 days with 95 percent of people having durations between 1.0 and 12.0 days. Notably, this implies that the incubation period can differ in duration from that of the latent period. We use the incubation duration to inform the rates of change from lower severity classes to higher severity classes (i.e., rates of transition from being asymptomatic to developing symptoms $\left(r_{1,a}^{E^j}, r_{1,a}^{I^k}\right)$). By examining the cumulative density function for the gamma distribution of incubation duration, we can determine the fraction of people at the beginning of each day since infection who are still asymptomatic and the daily change in the cumulative fraction of people who become symptomatic. These can be used to compute a daily rate of becoming symptomatic from the CDF which is initially accelerating, rising from 0.024 on the first day to 0.106 on the second, 0.186 on the third, 0.248 on the fourth, 0.296 on the fifth, etc..

We note that in the absence of mass population screening or random population screening, it is likely that symptomatic individuals (i.e., those in higher severity classes, $l = 2$) are more likely to be diagnosed and hence diagnosis rates ($\nu$) depend on an individual's symptom status (described in detail in the sections below). While the model also allows for the possibility of diagnosis via active screening ($\phi$), generally we set this rate to 0 for the historic periods to which we are calibrating.

We continue to actively review the literature to further update these parameter values as well as their uncertainties.

##### 1.2.2 Demographic parameters

To model a given population, the main groups of demographic parameters required include: 1) the total size of the population; 2) the age structure of the population (fraction of the total population in each age group); 3) the crude birth rate; 4) the age-specific background mortality rates (i.e., mortality due to all causes other than COVID-19); and 5) the ingredients to compute adjusted population density (total land area, urban land area, and fraction of total population living in urban areas). All of the above groups of demographic parameters are obtainable from public sources for many national and subnational geographic areas (i.e., counties of the United States or states of Mexico). However, adjusted population density deserves specific attention here because of its role in the model inputs and therefore how we specifically compute it.

For our modeling purposes, we are interested in population density because we wish to reflect the expected intensity of mixing between people (contacts per unit time) with higher density implying more intense mixing (discussed in further detail in the section on contact matrices below). Because of this, we opt for an alternative definition to those typically used for population density. Instead of defining population density as total population divided by the total area of a given jurisdiction or even the total population divided by the total land area (excluding water and other uninhabitable places), we focus on adjusted (weighted) population density.

Adjusted population density depends on the following inputs: land area and the fraction of the population living in urban areas. Land area (as opposed to total area) is important because we want to know how many people there are per unit area in potentially livable places. Fraction urban is important because urban areas typically occupy a small fraction of the total land area but can contain a high fraction of the population leading to very high densities (e.g., New York City has a population density in excess of 50,000 people per mile$^2$). Typically we would then assume that a relatively small number of square miles houses the fraction of the population that is urban multiplied by the total population yielding a high density and that the remaining rural population's average density is its size divided by the remaining land area. We then either model urban and rural populations separately or else take the average of their densities weighted by the fraction of the population that is urban/rural which will generally be higher than just dividing total population by the land area. To reflect relatively local differences, we try obtain such parameters at the finest geographic level possible which is typically county-level or state-level and likewise model at the finest possible geographic level.

While multiplying total population by percent urban yields the urban population, a challenge of this method for constructing the population-weighted density is knowing how much of the land area is urban land area. It is often possible to obtain lists of cities (e.g., for Brazil or Mexico) or urban and rural census tracts (e.g., for the U.S.) and their populations and land areas along with the state and/or county to which each city (or census tract) pertains. One can then compute the density of the urban population in each of the geographic areas and likewise the rural density by subtracting the urban land area from the total land area and the urban population from the total population. With these, one can then compute a population weighted average based on the relative sizes of the urban and rural populations.

For our applied analyses, we have obtained demographic parameters from publicly available sources: For example, for counties and states in the United States, we use: county population size and age structure from 2018;[10] state-specific life tables;[11] and county urban/rural status, land area, and population density.[12] Other specific countries on which we are implementing the model have their own sub-national sources. For other countries at a national level we use.[13,14]

##### 1.2.3 Estimation of contact matrices

To model potential transmission between subgroups, we use a contact (WAIFW) matrix approach. Entries in our contact matrix are the number of daily sufficient contacts a person in a given age group has with people of each age group in the model. A sufficient contact is defined as one that is close, long, and/or intense enough so that transmission could occur if one of the individuals was infectious and the other susceptible.

A data challenge for many sub-national populations is that there are no contact matrices estimated based on empirical data sampled from them; hence we develop an approximation method based on available data to estimate contact matrices. Specifically, we use publicly available national-level contact matrix estimates[15] along with epidemiological theory on how contact frequency/intensity depends on density in terms of functional form based on studies from several other human and animal diseases.[16,17]

We start with Prem et al.'s estimates of 152 national-level household and non-household contact matrices (total minus household contact matrices), $W^v$, whose structure is defined in (3), we compute average national-level age-weighted (based on population age structure) household and non-household contact rates defined as

$$rate^v_{contacts,weighted} = \sum_{a=1}^{N} p_a \left( \sum_{a'=1}^{N} W^v_{a,a'} \right). \tag{12}$$

Each entry in the matrix $W^v$ represents the number of venue-specific contacts per day by a person of age group $a$ with people of age group $a'$. We sum the number of contacts that each age group $a$ has over contacts for all the age groups and then compute the weighted average with weights equal to $p_a$, the proportion of the population in age group $a$.

As epidemiological theory suggests that contacts are related to population density, we estimate this relationship using a regression approach. We determine the population density of each country based on its urban population, urban land area, rural population, and rural land area, using these to compute the urban and rural population densities and the population-weighted average density.[18] We then take the natural logarithm of these densities for the regression. We regress the non-household contact rates on the logged population-distribution weighted density, to establish empirical estimates of the concave relationship for non-household contacts and no such relationship for household contacts per theorized relationships as described by Dalziel et al and Hu et al.[16,17]

The results of this regression provide us with two coefficients (the intercept and the slope for the log-density) which enable us to predict expected weighted average contacts for a population with any density. Our method makes use of the slope coefficient for the log-density.

For each of the 152 countries for which we have data, we employ the following procedure to form sub-national non-household contact matrix estimates (e.g., county-specific contact matrix estimates in the U.S. or state-specific contact matrix estimates in Mexico).

First, for each sub-national geography, we determine the population average weighted density which we transform using the natural logarithm. Next, we define a country-specific prediction equation using the slope coefficient from our regression above and determining the intercept based upon passing through the national-level population age-weighted non-household contact rate at the national-level log-density. With this prediction equation and the sub-national logged densities we generate sub-national predicted non-household contact rates. We take the ratio of the sub-national rates to the national rates to produce a regression-predicted density adjustment factor for each sub-national geography.

In addition to these density adjustment factors, in order to produce sub-national non-household contact matrices, we require a representation of the country's non-household contact matrix that is independent of its population age structure (a homogeneously mixing population whose subgroups are not all of equal size will have more people mixing with the more prevalent subgroups even without any assortative preference). To remove the age structure, we divide each age group's vector of age-specific contact rates (the $W^v_{a,a'}$s for each $a$ and for all $a'$s) by the proportion of the population in each age group ($p'_a$).

For each sub-national geography, we then multiply the age structure-removed non-household country-level contact matrix by the corresponding sub-national regression-predicted density adjustment factor; and finally, we multiply the population proportions from the sub-national area (its $p'_a$s) by the entries in the matrix to compute a matrix of appropriate contact frequency with the same underlying assortative preferences for between-group contacts in a population of the sub-national area's age structure.

Since the SC-COSMO model considers several different who mixes with whom matrices for non-household contacts (current venues include school, work, and other), the extension to the method above that we use in practice is that once we have computed the regression-predicted density adjustment factors for each sub-national geography, we apply them separately to each of the venue-specific national non-household contact matrices (after first removing the national population age structure from these matrices). We then apply the sub-national population age structure to all of the resulting matrices. This yields a set of venue-specific sub-national non-household contact matrices that depend on the sub-national geography's weighted population density and population age structure.

We presented further details of this approach at the 2020 Society for Medical Decision Making Annual Meeting.[19]

###### 1.2.4 Excess Mortality and Case Fatality Rates

Excess mortality risk due to COVID-19 infection (the Infection Fatality Rate [IFR]) is difficult to determine with currently available data because both the population at risk (i.e., denominator) and the number of observed COVID-19 deaths are limited to those individuals who are diagnosed with COVID-19. What can be computed directly from these observed quantities is the Case Fatality Rate (CFR) and its extension, the age-specific CFR. However, the CFR and its age-specific versions are likely overestimates of the corresponding IFRs in many situations. The reasons for this are several. First, CFRs do not include undiagnosed COVID-19 infections and deaths. Second, diagnosis without active surveillance selects for more severe cases which are more likely to die. Below we describe our current approach for quantifying deaths from COVID-19.

Furthermore, the overall CFR (or IFR) estimated in one population is also likely to be a biased estimate for other populations if their population age structures – specifically the age structures of COVID-19 cases/infections – differ substantially. This is because the overall CFR (or IFR) is a weighted mean where the weights are proportion of people in each age group at risk. Hence, we prefer the age-specific versions of these quantities, especially when transferring them from one population to another. An additional source of potential bias is that supportive medical care may modify the age-specific probabilities of death from COVID-19, and in some settings (e.g., low resource settings vs. wealthier settings) such differences could also be substantial. While we currently do not have a formal method for correcting for this sort of potential bias, we imagine an approach that incorporates measures/proxies of healthcare system efficacy relevant for conditions like COVID-19 and perhaps differences in mortality for other conditions (e.g., hospitalized pneumonia in general).

Our goal is to generate a set of age-specific COVID-19 mortality rates that are consistent with what has been described in the literature and how it is estimated, consistent with time-series of observed COVID-19 deaths in the jurisdictions we are modeling, and which is robust to potential changes in detection rates. We examine a variety of published sources on IFR and CFR including[20] along with the case series and death series from jurisdictions like counties in California and states in Mexico.

Additionally, aside from the fact that diagnosed individuals may receive supportive medical treatment, we want to ensure that the likelihood of death for any given individual is the same regardless of diagnosis status – that is, that if we randomly tested 10% versus if we randomly tested 20% of the population and examined the fractions of people who are infected with COVID-19 who die under each testing scenario, they should be the same. In reality, in many settings, testing is not done on random samples of the population and very likely is concentrated in individuals with more acute illness, those of certain ages, and other characteristics. For this reason, we allow testing rates to differ by such characteristics and over time. Most importantly for our explanation here, we allow testing (and hence diagnosis) to differ by severity level of illness and within a given severity level assume that death rates would be equal for individuals who are detected or not detected (if effective, supportive medical care were not given).

Currently the model assumes that for people with COVID-19 infections and no symptoms (i.e., severity level 1), the excess risk of death from COVID-19 is 0 (i.e., $CFR_{l=1,a} = 0$). For individuals who have developed symptomatic infections, there is an excess risk of death from COVID-19 ($CFR_{l=2,a} > 0$) which is not currently strongly modified by supportive medical care ($\alpha_{l=2}^{DX} = 1$) and that excess death risks have declined in time as testing and detection rates of somewhat less severe cases within severity level 2 have increased. When detailed and comprehensive data on the timing and mortality status of cases is available (as is the case in Mexico City Metropolitan Area and other states in Mexico), we estimate time-varying case fatality rates using statistical regressions. In other cases, we calibrate these excess death rates to observed, jurisdiction-specific time-series on COVID-19 deaths.

For settings like the United States, the CDC and other departments of health have published outcomes including case fatality rates for an initial cohort of individuals as well as other information suggesting infection fatality rates, and additional data are increasingly becoming available in the published literature as well as through state reporting. We will continue to gather information to update excess mortality risks as new data become available.

###### 1.2.5 Case detection rate

Just like fatality rates, detection rates are challenging to estimate because we lack an important component of the denominator, the total number of prevalent infections that could be detected if perfectly sensitive and specific testing were applied to all of them. The metrics that are more commonly reported are: 1) the time-series of total tests performed; 2) the time-series of the fraction of people tested who are positive for COVID-19. But the relationship between the commonly reported metrics and the detection rate is likely time-varying and confounded by a number of other factors.

To begin to get a sense of the possible confounding and complexity, one can imagine the process (ignoring age groups) as the following. At a given point in time, people who are infected with COVID-19 have a range of symptoms/severity. All else equal, we would expect people with more severe symptoms to seek testing more frequently. However, we would also expect people with other Influenza-like Illnesses (ILIs) or who may believe they were exposed to COVID-19 to also be more likely to seek testing. For both groups, those who seek testing/care will interact with the healthcare system. At a given point in time, the system has specific criteria for testing (which may or may not be strictly followed) as well as a supply constraint on the number of tests they can perform. So the number of potentially true and false positive individuals who are tested depends on these factors and hence the case detection rate is likely time-varying and challenging to determine.

We have described our approach to putting bounds on the detection rate based upon logical constraints and available data on the testing capacity time-series and other modeling, which was presented at the 2020 Society for Medical Decision Making Annual Meeting.[21]

###### 1.2.6 Risk of hospitalization and conditional risk of requiring ICU given hospitalization

Risks of hospitalization depend on setting. For example, Verity et al. 2020[20] report risks of hospitalization conditional on infection for COVID-19 patients in China which can differ from those in Mexico or in the United States. Because of this, for specific geographic areas/populations whose healthcare system characteristics may differ, we try to incorporate local data whenever they are available and of sufficiently high quality. It is possible to estimate probabilities and other quantities required by the hospitalization module directly from data that is sufficiently detailed as we do with data from Mexico City Metropolitan Area. For remaining quantities (potentially time-varying) required by the hospitalization module, it is possible to calibrate them based on time-series of hospitalized prevalence (e.g., daily COVID-19 bed census).

For detected COVID-19 positive individuals who are hospitalized, some fraction will ultimately have more severe illness than others. Depending on illness severity, hospitalized patients have a risk of being placed in the intensive care unit (ICU) or requiring a ventilator. Depending on severity and ICU or ventilator status, patients have different length of stay distributions. Hospitalized COVID-19 prevalence at any given point in time is therefore determined by the number of incident cases entering the hospital each day prior to this point in time and the fraction of those people remaining in the hospital for a sufficient length of time such that they have not yet exited (i.e., leaving due to death or discharge):

$$Hosp(T) = \sum_{a} \left( \sum_{t \leq T} IncDX_{a,t} * pHosp_{a,t} \left( \sum_{s,i} prop_{a,s,i}(1 - CDF_{a,s,i}(T - t)) \right) \right) \tag{13}$$

where $T$ is the date for which we are interested in knowing the census of hospitalized COVID-19 patients ($Hosp$); $t$ is a date that happens on or before $T$; $a$ is an age group; $IncDX_{a,t}$ is the number of individuals with incident diagnoses of COVID-19 for each age group on each date; $pHosp_{a,t}$ is the probability of hospitalization among individuals with diagnosed COVID-19 of a given age group diagnosed on a given date; $s$ indexes more or less severe hospitalized illness; $i$ indexes ICU and non-ICU treatment; $prop_{a,s,i}$ is the proportion of hospitalized COVID-19 patients of a given age group whose illness is more or less severe and who are or are not treated in the ICU; and $CDF_{a,s,i}$ is the cumulative density function of length of stay in the hospital for COVID-19 admitted patients conditional on age group, severity, and ICU status for which we use gamma distributions. For Mexico City Metropolitan Area, we extend this approach to have these length of stay distributions depend on the date at which hospitalization occurs.

From above, $IncDX_{a,t}$ is clearly specific to particular geographic areas. Likewise, the probabilities of hospitalization, the severity and ICU mixture of hospitalized patients, and the length of stays for each patient group likely vary with local geography. Reports on severity, ICU and length of stay include studies by Guan et al. 2020 and Lewnard et al. 2020 among others.[22,23] In brief, Guan et al. report the number of hospitalized patients within each of their age categories who are severe or non-severe. From these and similar sources, we are able to compute quantities like: the age-specific probability a hospitalized patient was severe; the probability of requiring ICU care conditional on severity (which does not appear to be strongly conditional on age once severity is taken into consideration); and length of stay distributions overall by age as well as distributions of additional time spent in the ICU. These quantities can inform the parameters in the model above (e.g., length of stay estimates inform the CDFs; conditional probabilities of severity inform the proportions facing different lengths of stay; etc.) when direct jurisdiction-specific quantities are unavailable. The likelihood of hospitalization conditional on detection is observed in some data sets (e.g., states of Mexico) but must be calibrated (e.g., counties in California).

###### 1.2.7 Interventions to reduce transmission

The model currently includes non-pharmaceutical interventions (NPIs) which represent various forms of social distancing and masking (i.e., they act to reduce contacts and/or the probability of transmission given contacts). The size of the intervention's effect can vary across age group. Interventions have start and end times. Time-varying intervention effects can therefore be constructed by defining a series of interventions whose start and end times bookend one another and whose effect sizes can be different from one another. For example, an intervention may have a strong effect at the time when shelter-in-place orders were implemented but over time with re-openings its effect may attenuate somewhat depending on continuing compliance with mask orders.

In general, modeled interventions can reduce effective contacts differentialy by venue. The intervention has no effect on within-home contacts between household members. The intervention reduces contacts at work and other venues proxying a variety of changes: e.g., greater distances and masking in outdoor locations like parks, more work-from-home, business conducted outside, masking, and limits on the percentage capacity at which in-person businesses are allowed to operate. Modeled interventions have two variants with respect to school contacts. The intervention can be accompanied by school closures in which case there are no school contacts or else it can allow return to school with some level of resumed contacts relative to pre-COVID school contact levels.

Another category of intervention effect is how well isolation of detected cases can occur. As noted above, the model allows for a parameter that reduces contact/transmission for detected infectious individuals.

A final intervention category includes vaccines, of which a number of candidates are currently in phase 3 trials but none have been approved to date. Current implementation work is extending the model's capability to include vaccination. This implementation work includes a vaccinated compartment in the model, creating realistic timing and targeting of vaccination coverage scale-up, allowing for partial effectiveness and potentially waning vaccine immunity which may differ from naturally acquired immunity, and incorporating vaccination effects within the household submodel.

Parameterization of the NPI interventions is challenging because there are currently no direct measures on the effect of interventions on effective contact frequencies, duration, or intensity. A variety of sources are attempting to measure parts or proxies of this. The simplest are surveys of people asking them about how they have changed a variety of their activities, how much time they spend in their homes, and for what reasons they go out. Simple indirect measures include quantities like the amount of air population measured by area which shows declines in commuter traffic and other production that yields emissions but does not show which types of trips are being curtailed. More sophisticated measures focus on contacts (e.g., the collocation of cellphones based on triangulation from towers or the location information of devices running various apps or the temporal proximity of credit card purchases by different purchasers at the same store) or on mobility with or without collocation (e.g., the fraction of time that someone's device is not in the location it is during typical sleeping hours or the number of devices on public transport, or how far devices travel away from home). Average changes in these measures compared to pre-COVID levels either by individual or by geography all form estimates of components of reductions in contacts sufficient to transmit COVID. For many of these measures, at least in California counties, there are average reductions of 10-70 percent or more which differ by county. What has not been reported to date are measures of individual (or small area) variance of such reductions within county, which are likely important for considering potential transmission among subgroups who must go out of their home (e.g., essential services providers) or who do not comply and then secondary transmission from these groups. Furthermore, when people are mobile or collocate it is not clear from these measures whether they are taking more or less precautions such as masking.

We currently use a composite approach to determine the timing and effect size of interventions by geographic location. This involves extracting needed information from mobility data and model calibration (described below). Currently we use multi-location mobility time-series data from sources like Google Mobility Trends and FourSquare to determine when, after NPIs were first implemented, these time-series change. For example, directly after NPI implementation when lock-downs were often in effect, the amount of travel to work locations dropped dramatically but that effect attenuated over the next month or so and then with re-openings and second viral waves may have fluctuated. By analyzing location-specific mobility time-series we determine when there are change-points for the modeled intervention intensity in a given geographic location. However, because changes in mobility patterns do not necessarily correspond to the full range of changes in contacts nor to how people may increase mask use for the same number of contacts, we use model calibration to incidence data to inform the effect sizes for each segment of the time-varying, geography-specific intervention effects. Additionally, we use and have recently made open source and public a dataset containing county-specific information on public health orders, categorized by the type of activity(s) they pertain to and the level of strictness.[24]

### 2 Calibration

A challenge for many mathematical simulation models is that they may require input values that are unobserved or unobservable due to financial, practical, or ethical reasons. In such situations, model calibration can be used to infer these values. Specifically, calibration is the process of estimating values for unknown or uncertain parameters of a mathematical model by matching its outputs to observed clinical or epidemiological data (known as calibration targets). The general goal of calibration is to identify set(s) of parameter values that maximize the fit between model outputs and the calibration targets.[25,26]

For the SC-COSMO model, calibration is required for a number of parameters that are not directly observed. These include the community transmission rates ($\beta_a^k$), the household transmission rate ($\tau$), the time-varying case detection rates $\left(\nu_l^{E^j} \text{ and } \nu_l^{I^k}\right)$, and the time-varying, venue-specific effects of interventions on community transmission rates ($\kappa_t^v$). Generally priors on parameters are defined based on existing knowledge about their values.[27] As there is substantial uncertainty in these input parameters, we employ uniform prior distributions for each. The prior distributions are uncorrelated, and they are relatively wide to reflect the underlying uncertainty. The prior ranges are defined for particular settings (i.e., the prior ranges for counties in California may differ from those used for the states of Mexico).

Calibration targets are formed by time-series of incident detected case counts both prior to and after the implementation of NPIs. As the targets are counts, we construct a likelihood function by assuming that the calibration targets ($y_{i,t}$) are Negative Binomial distributed

$$y_{i,t} \sim \text{NegBin}\left(\mu_{i,t}, size_i\right) \tag{14}$$

where $\mu_{i,t}$ is the model-predicted output for each type of target $i$ (e.g., case count) at each time $t$ and $size_i$ is the dispersion parameter such that

$$\begin{aligned} prob_{i,t} &= size_i / \left(size_i | \mu_{i,t}\right) \\ variance_{i,t} &= \mu_{i,t} + \left(\mu_{i,t}^2 / size_i\right) \end{aligned} \tag{15}$$

per alternative parameterizations of the distribution.

We use two main algorithms to find set(s) of input parameters that cause the model's outputs to match the calibration targets. The first is the incremental mixture importance sampling (IMIS) algorithm,[28,29] which has been previously used to calibrate deterministic health policy models.[30–32] An advantage of IMIS over other Monte Carlo methods, such as Markov chain Monte Carlo, is that with IMIS the evaluation of the likelihood for different sampled parameter sets can be parallelized, which makes its implementation especially suitable for an high-performance computing (HPC) environment such as the ones used for SC-COSMO calibration. The second is the Nelder-Mead (NM) algorithm.[33] This directed search approach is efficient at identifying critical points with complex models because it is gradient-free and guarantees convergence to locally optimal solutions. To identify multiple sets of parameters that are consistent with the calibration targets, we start the NM-based calibration from multiple random starting locations within our prior space. For both algorithms, when we are only interested in identifying a single, best-fit parameter set without running the sampling algorithm, we adopt a Laplace approximation where we compute the posterior mode often called the maximum a posteriori (MAP) estimate, by maximizing the logarithm of the posterior, and use the MAP estimate (instead of the mean) as an approximation of the parameter set $\theta$. The inverse of the negative Hessian of the logarithm of the posterior can be used to measure the uncertainty of this approximation.[34–38] In the case of the SC-COSMO model, we use not only the mean and hence use the calibrated posterior parameter distributions for projections and analyses of scenarios.

### 3 Outcomes of interest

All the compartments in our models are variables that depend on time, $t$; however, we omit this index to simplify the notation. We also omit the index, $l$, referring to severity levels of exposed and infectious compartments as well as diagnosed exposed and infectious compartments.

#### 3.1 Demographic outcomes

##### 3.1.1 Population

The age-group-specific population, $Pop_a$, is given by

$$Pop_a = S_a + \sum_{j=1}^{J} E_a^j + \sum_{j=1}^{J} EDX_a^j + \sum_{k=1}^{K} I_a^k + \sum_{k=1}^{K} IDX_a^k + R_a, \tag{16}$$

and the total population across all age groups is given by

$$Pop = \sum_{a=1}^{N} Pop_a. \tag{17}$$

#### 3.2 Epidemiological outcomes

##### 3.2.1 Cumulative infectious individuals

The age-group-specific cumulative numbers of infectious individuals, $CI_a$, are given by

$$CI_a = \int_0^T \sigma J \left( E_a^J + EDX_a^J \right) dt, \tag{18}$$

and the total cumulative numbers of infectious individuals across all age groups is given by

$$CI = \int_0^T \sigma J \left( \sum_{a=1}^{N} \left( E_a^J + EDX_a^J \right) \right) dt, \tag{19}$$

where $T$ is the analytic horizon.

###### 3.2.2 Infectious individuals

The age-group-specific total numbers of infectious individuals at any time $t$, $TotI_a$, are given by

$$TotI_a = \sum_{k=1}^{K} \left(I_a^k + IDX_a^k\right), \tag{20}$$

and the total number of infectious individuals across all age groups is given by

$$TotI = \sum_{a=1}^{N} TotI_a, \tag{21}$$

###### 3.2.3 Incident infectious individuals

The age-group-specific incident numbers of infectious individuals, $IncI_a$, are given by

$$IncI_a = \sigma J \left(E_a^J + EDX_a^J\right), \tag{22}$$

and the total incident infections across all age groups is given by

$$IncI = \sum_{a=1}^{N} IncI_a. \tag{23}$$

###### 3.2.4 Cumulative diagnosed infections (regardless of infectiousness)

The age-group-specific cumulative diagnosed infections, $CIDX_a$, are given by

$$CDX_a = \int_0^T \left( \sum_{j=1}^{J} \left(\left(\nu^j + \phi^j\right) E_a^j\right) + \sum_{k=1}^{K} \left(\left(\nu^k + \phi^k\right) I_a^k\right) \right) dt, \tag{24}$$

and the total cumulative diagnosed infections across all age groups is given by

$$CDX = \int_0^T \sum_{a=1}^{N} \left( \sum_{j=1}^{J} \left(\left(\nu^j + \phi^j\right) E_a^j\right) + \sum_{k=1}^{K} \left(\left(\nu^k + \phi^k\right) I_a^k\right) \right) dt, \tag{25}$$

where $T$ is the analytic horizon.

###### 3.2.5 Diagnosed Infections (regardless of infectiousness)

The age-group-specific total numbers of diagnosed infections at any time $t$, $DX_a$, are given by

$$DX_a = \sum_{j=1}^{J} EDX_a^j + \sum_{k=1}^{K} IDX_a^k, \tag{26}$$

and the total diagnosed infections across all age groups is given by

$$DX = \sum_{a=1}^{N} DX_a. \tag{27}$$

##### 3.2.6 Incident diagnosed infections (regardless of infectiousness)

The age-group-specific numbers of incident diagnosed infections, $IncDX_a$, are given by

$$IncDX_a = \left( \sum_{j=1}^{J} \left( \left( \nu^j + \phi^j \right) E_a^j \right) + \sum_{k=1}^{K} \left( \left( \nu^k + \phi^k \right) I_a^k \right) \right), \tag{28}$$

and the total incident diagnosed infections across all age groups is given by

$$IncIDX = \sum_{a=1}^{N} IncIDX_a. \tag{29}$$

##### 3.2.7 Total COVID-19 deaths

The age-group-specific total COVID-19 deaths, $TotDCOVID_a$, are given by

$$TotDCOVID_a = \int_0^T \left( p_l^d \gamma K \left( I_{l,a}^K \right) + \alpha_l^{DX} p_l^d \gamma K \left( IDX_{l,a}^K \right) \right) dt, \tag{30}$$

and the total cumulative COVID-19 deaths across all age groups is given by

$$TotDCOVID = \int_0^T \sum_{a=1}^{N} \left( p_l^d \gamma K \left( I_{l,a}^K \right) + \alpha_l^{DX} p_l^d \gamma K \left( IDX_{l,a}^K \right) \right) dt, \tag{31}$$

where $T$ is the analytic horizon.

##### 3.2.8 Known COVID-19 deaths

The known cumulative COVID-19 deaths are those observed from diagnosed infected cases only. The age-group-specific known COVID-19 deaths, $KnownDCOVID_a$, are given by

$$KnownDCOVID_a = \int_0^T \alpha_l^{DX} p_l^d \gamma K \left( IDX_{l,a}^K \right) dt, \tag{32}$$

and the total known COVID-19 deaths across all age groups is given by

$$KnownDCOVID = \int_0^T \sum_{a=1}^{N} \alpha_l^{DX} p_l^d \gamma K \left( IDX_{l,a}^K \right) dt, \tag{33}$$

where $T$ is the analytic horizon.

### References


[1] Matthew James Keeling and Pejman Rohani. *Modeling Infectious Diseases in Humans and Animals*. Princeton University Press, Princeton, N.J., 2008.

[2] Lorenzo Pellis, Simon Cauchemez, Feruson Neil M., and Fraser Christophe. Systematic selection between age and household structure for models aimed at emerging epidemic predictions. *Nature Communications*, 11(1), 2020.

[3] Thomas House and Matt J. Keeling. Deterministic epidemic models with explicit household structure. *Mathematical Biosciences*, 213(1):29 – 39, 2008.

[4] Hannah F. Fung, Leonardo Martinez, Fernando Alarid-Escudero, Joshua A. Salomon, David M. Studdert, Jason R. Andrews, Jeremy D. Goldhaber-Fiebert, and SC-COSMO Modeling Group. The household secondary attack rate of SARS-CoV-2: A rapid review. *Clinical Infectious Diseases*, 2020.

[5] S Y Del Valle, J M Hyman, H W Hethcote, and S G Eubank. Mixing patterns between age groups in social networks. *Social Networks*, 29:539–554, 2007.

[6] Fernando Alarid-Escudero, Andrea Luviano, Natalia Kunst, Yadira Peralta, Jeremy D Goldhaber-Fiebert, and SC-COSMO Modeling Group. Accounting for Household Transmission Dynamics in Realistic Epidemic Models [top-rated abstract]. In *The 42nd Annual Meeting of the Society for Medical Decision Making*, 2020.

[7] Stephen A. Lauer, Kyra H. Grantz, Qifang Bi, Forrest K. Jones, Qulu Zheng, Hannah R. Meredith, Andrew S. Azman, Nicholas G. Reich, and Justin Lessler. The Incubation Period of Coronavirus Disease 2019 (COVID-19) From Publicly Reported Confirmed Cases: Estimation and Application. *Annals of Internal Medicine*, 03 2020.

[8] Xi He, Eric H. Y. Lau, Peng Wu, Xilong Deng, Jian Wang, Xinxin Hao, Yiu Chung Lau, Jessica Y. Wong, Yujuan Guan, Xinghua Tan, and et al. Temporal dynamics in viral shedding and transmissibility of covid-19. *Nature Medicine*, Apr 2020.

[9] P. Ashcroft, J. S. Huisman, S. Lehtinen, J. A. Bouman, C. L. Althaus, R. R. Regoes, and S. Bonhoeffer. Covid-19 infectivity profile correction. *Swiss medical weekly*, 150(w20336), 2020.

[10] US Census Bureau. County population by characteristics: 2010-2018, Feb 2020.

[11] University of California Berkeley. United states mortality database, Feb 2020.

[12] Economic Research Service United States Department of Agriculture. Rural-urban commuting area codes, Oct 2019.

[13] United Nations Department of Economic and Population Dynamics Social Affairs. World population prospect 2019, quinquennial population by five-year age groups - both sexes. de facto population as of 1 july of the year indicated classified by five-year age groups (0-4, 5-9, 10-14, ..., 95-99, 100+). data are presented in thousands., Oct 2019.

[14] United Nations Department of Economic and Population Dynamics Social Affairs. World population prospect 2019, number of survivors by age for a hypothetical cohort of 100,000 newborns who would be subject during all their lives to the mortality rates of a given period., Oct 2019.

[15] Kiesha Prem, Alex R. Cook, and Mark Jit. Projecting social contact matrices in 152 countries using contact surveys and demographic data. *PLoS Computational Biology*, 13(9):e1005697, sep 2017.

[16] Benjamin D. Dalziel, Stephen Kissler, Julia R. Gog, Cecile Viboud, Ottar N. Bjørnstad, C. Jessica E. Metcalf, and Bryan T. Grenfell. Urbanization and humidity shape the intensity of influenza epidemics in u.s. cities. *Science*, 362(6410):75–79, 2018.

[17] Hao Hu, Karima Nigmatulina, and Philip Eckhoff. The scaling of contact rates with population density for the infectious disease models. *Mathematical Biosciences*, 244(2):125 – 134, 2013.

[18] Center for International Earth Science Information Network CIESIN Columbia University. NASA Socioeconomic Data and Applications Center (SEDAC). Low elevation coastal zone (lecz) urban-rural population and land area estimates, version 2, 2013.

[19] Marissa Reitsma, Fernando Alarid-Escudero, Jason Andrews, David Studdert, Natalia Kunst, Anneke Claypool, Tess Ryckman, Jeremy D. Goldhaber-Fiebert, and SC-COSMO Modeling Group. Methods for Constructing Sub-National Contact Matrices for Transmission Models of Respiratory Viruses like SARS-Cov-2 (COVID-19)[abstract]. In *The 42nd Annual Meeting of the Society for Medical Decision Making*, 2020.

[20] Robert Verity, Lucy C Okell, Ilaria Dorigatti, Peter Winskill, Charles Whittaker, Natsuko Imai, Gina Cuomo-dannenburg, Hayley Thompson, Patrick G T Walker, Han Fu, Amy Dighe, Jamie T Griffin, Marc Baguelin, Sangeeta Bhatia, Adhiratha Boonyasiri, Anne Cori, Zulma Cucunubá, Rich Fitzjohn, Katy Gaythorpe, Will Green, Arran Hamlet, Wes Hinsley, Daniel Laydon, Gemma Nedjati-gilani, Steven Riley, Sabine Van Elsland, Erik Volz, Haowei

Wang, Yuanrong Wang, Xiaoyue Xi, Christl A Donnelly, Azra C Ghani, and Neil M Ferguson. Estimates of the severity of coronavirus disease 2019: a model-based analysis. *Lancet Infectious Diseases*, 3099(20):1–9, 2020.

[21] Tess Ryckman, Fernando Alarid-Escudero, Natalia Kunst, Jeremy D Goldhaber-fiebert, and SC-COSMO Modeling Group. A New Method for Bounding the Case Detection Fraction of an Emerging Epidemic and a COVID-19 Application [abstract]. In *The 42nd Annual Meeting of the Society for Medical Decision Making*, 2020.

[22] Wei-jie Guan, Zheng-yi Ni, Yu Hu, Wen-hua Liang, Chun-quan Ou, Jian-xing He, Lei Liu, Hong Shan, Chun-liang Lei, David S.C. Hui, Bin Du, Lan-juan Li, Guang Zeng, Kwok-Yung Yuen, Ru-chong Chen, Chun-li Tang, Tao Wang, Ping-yan Chen, Jie Xiang, Shi-yue Li, Jin-lin Wang, Zi-jing Liang, Yi-xiang Peng, Li Wei, Yong Liu, Ya-hua Hu, Peng Peng, Jian-ming Wang, Ji-yang Liu, Zhong Chen, Gang Li, Zhi-jian Zheng, Shao-qin Qiu, Jie Luo, Chang-jiang Ye, Shao-yong Zhu, and Nan-shan Zhong. Clinical Characteristics of Coronavirus Disease 2019 in China. *New England Journal of Medicine*, pages 1–13, 2020.

[23] Joseph A Lewnard, Vincent X Liu, Michael L Jackson, Mark A Schmidt, Britta L Jewell, Jean P Flores, Chris Jentz, Graham R Northrup, Ayesha Mahmud, Arthur L Reingold, Maya Petersen, Nicholas P Jewell, Scott Young, and Jim Bellows. Incidence, clinical outcomes, and transmission dynamics of severe coronavirus disease 2019 in california and washington: prospective cohort study. *BMJ*, 369, 2020.

[24] Jeremy D. Goldhaber-Fiebert, Alex Holsinger, Erin Holsinger, Elizabeth Long, and SC-COSMO Modeling Group. State- and County-Level COVID-19 Public Health Orders in California: Constructing a Dataset and Describing Their Timing, Content, and Stricture. 2020.

[35] Bradley P. Carlin and Thomas A. Louis. *Bayesian Methods for Data Analysis*. CRC Press, third edition, 2009.

[36] Tan Bui-Thanh, Omar Ghattas, James Martin, and Georg Stadler. A Computational Framework for Infinite-Dimensional Bayesian Inverse Problems Part I: The Linearized Case, with Application to Global Seismic Inversion. *SIAM Journal on Scientific Computing*, 35(6):2494–2523, 2013.

[37] Marco A. Iglesias, Kody J H Law, and Andrew M. Stuart. Evaluation of Gaussian approximations for data assimilation in reservoir models. *Computational Geosciences*, 17(5):851–885, 2013.

[38] Matthias Morzfeld, Xuemin Tu, Jon Wilkening, and Alexandre J. Chorin. Parameter estimation by implicit sampling. *Communications in Applied Mathematics and Computational Science*, 10(2):205–225, 2015.
